# Individual, organisational and system capacities, and the functioning of a multi-country implementation-focused network for maternal, newborn and child health: Bangladesh, Ethiopia, Malawi, and Uganda

**DOI:** 10.1101/2023.03.04.23286667

**Authors:** Anene Tesfa, Catherine Nakidde, Kohenour Akter, Fatama Khatun, Kondwani Mwandira, Seblewengel Lemma, Gloria Seruwagi, Kasonde Mwaba, Mike English, Callie Daniels-Howell, QCN Evaluation Group, Nehla Djellouli, Tim Colbourn, Tanya Marchant

**Affiliations:** Ethiopian Public Health Institute, Addis Ababa, Ethiopia; School of Public Health, Makerere University, Kampala, Uganda; Perinatal Care Project, Diabetic Association of Bangladesh, Dhaka, Bangladesh; Parent and Child Health Initiative PACHI, Lilongwe, Malawi; Department of Disease Control, London School of Hygiene & Tropical Medicine, London, UK; Institute for Global Health, University College, London, UK; Centre for Tropical Medicine and Global Health, University of Oxford, Oxford, UK

**Keywords:** capacities, multi-county network, quality of care, maternal and newborn health, Bangladesh, Ethiopia, Malawi, Uganda

## Abstract

Better policies, investments, and programs are needed to improve the integration and quality of maternal, newborn, and child health services. Previously, partnerships and collaborations that involved multiple countries with a unified aim have been observed to yield positive results. Since 2017, the WHO and partners have hosted the Quality of Care Network [QCN], a multi-country implementation network focused on improving maternal, neonatal, and child health care. In this paper we examine the functionality of QCN in different contexts. We focus on implementation capacities and contexts in four network countries: Bangladesh, Ethiopia, Malawi, and Uganda. In each country, the study was conducted over several consecutive rounds between 2019-2022, employing 227 key informant interviews with major stakeholders and members of the network countries, and 42 facility observations. The collected data were coded using Nvivo-12 software and categorized thematically. The study showed that individual, organizational and system-level capacities, and circumstances all played an important role in shaping implementation success in network countries, but that these levels were inter-linked. Across all levels, systems that enabled leadership, motivated and trained staff, and created a positive culture of data use were critical – from the policy making arena including addressing financing issues - to the day-to-day practice improvement at the front line. Some characteristics of QCN actively supported these levels, for example shared learning forums for continuous learning, a focus on data and tracking progress, and emphasising the importance of coordinated efforts towards a common goal. However, inadequate system financing and capacity also hampered network functioning, especially in the face of external shocks.

## Introduction

Despite advances in maternal and newborn survival rates in low and middle-income countries, most deaths are attributed to weak health systems and poor quality maternal and newborn healthcare services [1]. Many maternal and newborn deaths are preventable if all women have access to quality care during and after childbirth and measures taken to support maternal health will also positively influence newborn health [2,3].

Lessons learnt from previous efforts to improve maternal, newborn and child healthcare such as improvement in coverage and quality of services have pointed to the need for integrated, comprehensive, and holistic approaches and systems that promote an enabling environment to produce a tangible result. Such environments are influenced by a combination of the circumstances of individuals, organizations, and systems so that quality services with the requisite staffing and equipment are supported by local problem-solving initiatives and coherent policies [4-6]. For example, the current recommendation for all births to be assisted by skilled professionals who provide care with knowledge, problem-solving attitudes and compassion is important but not sufficient to save lives as these individuals also require an appropriate environment to practice in [7,8]. Therefore, at the organizational level, foundations such as the availability of basic amenities and adequate equipment and staff, favourable rules, and regulations as well as a supportive management system are needed to stimulate the implementation of interventions aimed at improving health [9]. And both individual and organizational performance requires system support [10].

One such initiative designed to take a holistic approach to service improvement is the network for improving Quality of Care for Maternal, Newborn and Child Health [QCN], a multi-country implementation and learning network focusing on maternal, newborn and child health care improvements, hosted by the World Health Organisation (WHO) since 2017 [11]. In 2017, the WHO and global partners launched QCN, with the partnership of ten pathfinder network countries: Bangladesh, Cote d’Ivoire, Ethiopia, Ghana, India, Malawi, Nigeria, Sierra Leone, the United Republic of Tanzania and Uganda. Kenya joined the network as the 11th country in 2019.

The launch of QCN presented an opportunity to learn lessons about this multi-country network strategy and implementation process for maternal and newborn health care quality improvement. This paper is part of a collection looking at different aspects of the emergence, legitimacy, and effectiveness of the network (S1 Text). Here, we report on the capacities of individuals, organizations, and systems that enabled or impeded the network’s functionality examining the implementation context in four of the network countries, Bangladesh, Ethiopia, Malawi, and Uganda.

## Methods

Our research provides the implementation perspective of these four network countries (S2 Text). The study teams in these countries used a mixed-method approach to collect data from the national and subnational level network members, including at the health facility level, for the overall evaluation. Data from qualitative interviews and facility observations are analysed and presented in this paper (Table 1).

**Table 1:** QCN interviews and observations completed.

| Case-study country | Data collection round (dates) | National interviews (n) | Local interviews (n) | Facility observation (n) |
| --- | --- | --- | --- | --- |
| Bangladesh | 1 (Oct 2019 – Mar 2020) | 13 | 07 | 3 |
|  | 2 (Oct 2020 – Jan 2021) | 14 | 11 | 4 |
|  | 3 (May 2021 - Sep 2021) | 10 | 12 | 4 |
|  | 4 (Jan 2022 - Mar 2022) | 08 | 00 | 00 |
| Ethiopia | 1 (Jan 2021– Mar 2021) | 08 | 11 | 4 |
|  | 2 (Nov 2021- Dec 2021) | 10 | 11 | 3 |
| Malawi | 1 (Oct 2019 – Mar 2020) | 07 | 12 | 4 |
|  | 2 (Nov 2020– Jan 2021) | 10 | 07 | 4 |
|  | 3 (Aug 2021 – Nov 2021) | 9 | 7 | 4 |
|  | 4 (Mar 2022) | 2 | 3 | 00 |
| Uganda | 1 (Nov 2020 – Mar 2021) | 07 | 13 | 4 |
|  | 2 (Jun 2021 – Sep 2021) | 12 | 08 | 4 |
|  | 3 (Feb 2022 – March- 2022) | 10 | 05 | 4 |

We conducted semi-structured interviews with national level and sub-national level network members and key stakeholders in Bangladesh, Ethiopia, Malawi, and Uganda. Participants included employees of government bodies, implementing and technical partners as well as health facility workers in the implementation districts. A topic guide was centrally prepared then translated to local languages (S2 Text) where appropriate and piloted in each country.

As a result, minor adaptations were made to reflect country contexts. In each country we conducted several rounds of interviews, at least six months apart, to capture changes in how the network was operating and views pertaining to network activities as well as follow-up on emerging findings from the previous round. Following the first round of data collection, topic guides were refined in subsequent rounds specifically for each country and category of stakeholders.

Facility observations were conducted in purposefully selected best and least well performing facilities in each country. These facilities were selected in conversation with network leaders in each country and based on health outcome data such as a comparison of maternal and child mortality indicators [available from national databases] and other quality of care data [e.g., those used in national schemes]. During observations, we used a common template to capture key processes relevant to the focus of the network in each country and recorded unstructured notes about health care processes on the day. Mothers coming to the facility were observed during service provision. Further details on our methods are provided in S2 Text.

Data were entered into Nvivo-12 software for analysis and all countries used a common codebook developed by the senior members of the QCN evaluation group, that was then iteratively refined to accommodate country-specific context (S2 text). We used a framework analysis for the purpose of writing this paper, informed by a conceptual framework for understanding policy competences and capabilities (figure 1) [12].

**Fig. 1.**
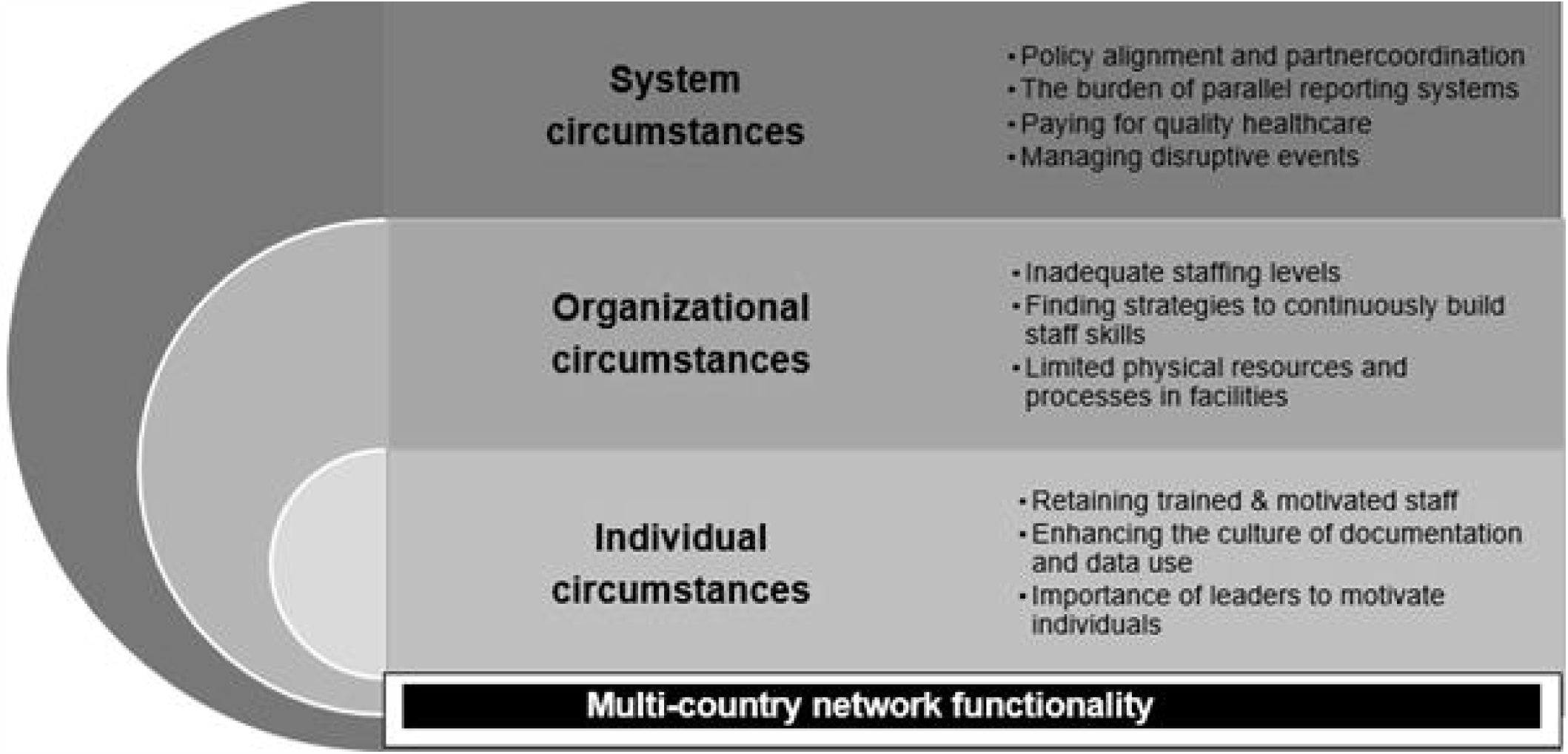
Conceptual framework on policy competencies and capabilities, adapted from Wu et. al [12].

### Ethical considerations

All interviews and observations were conducted by local team members after obtaining informed, formal written consent, including separate written consent for tape recording. Patients’ privacy was respected during hospital observations. All data are confidential and anonymised. Ethical approval was obtained from the Research Ethics Committee at University College London (3433/003) and institutional review boards in Bangladesh (BADAS-ERC/EC/19/00274), Ethiopia (EPHI-IRB-240-2020), Malawi (NHSRC Ref, 19/03/2264) and Uganda (MUSPHHDRC, Ref: 869).

## Results

Here we present findings on ways in which individual, organizational and system circumstances influenced the functionality of QCN at the national level, highlighting common themes to emerge across the four network countries.

### Individual circumstances

Three key themes emerged under individual circumstances: [i] the need to retain trained and motivated staff, and the frustration experienced when this was suboptimal; [ii] enhancing a culture of documentation and data use; and [iii] the importance of leaders to motivate individuals.

#### i. The need to retain trained and motivated staff

QCN was perceived to have improved the availability of skilled professionals in enrolled facilities. This is both by increasing the number of staff and improving the skills of the existing ones. Bangladesh is one of the countries making progress in consistently providing health care with the assistance of trained professionals. However, participants reported on-going challenges and frustrations in retaining quality improvement trained and motivated staff. In Bangladesh while facilities were striving to provide full-time skilled care, it was noted that not all staff provided an equally high-quality service and high demand negatively affected staff motivation.

> *“Clinical standards are not uniformly applied by all providers; there are insufficient training and decision aids; there are long patient waiting times and improper queue management and a large staff workload has resulted in challenges with maintaining staff motivation*.*” [QCN webinar-Global level-Bangladesh Round 1]*

Newly qualified and fresh staff joining the health system in Ethiopia and Uganda were thought to contribute to a lack of saturation of quality improvement (QI) knowledge and skills. And this QI skill gap was exacerbated by high levels of staff attrition, affecting the institutional memory for network functioning. In Malawi, retention of skilled staff was a significant challenge as well, partly due to a pre-existing ministry of health policy in place that rotates health care workers between departments and/or health facilities, thereby disrupting institutional memory about QI and network activities.

> *“Staff turnover due to rotation to other departments/facilities affects the quality of Care as health workers trained to provide MNH services are moved out and incoming health workers may not have the necessary skills. This also directly affects QCN implementation as sometimes the staff who are shifted are trained WIT [Work Improvement Team] members*.*” [Facility observation-Case 3-Malawi Round 2]*

One of the assumptions of the network was that knowledge and learning would be shared among network member health professionals and periodic learning forums were organized for this purpose. Staff attending these forums were eligible for an allowance to cover any expenses and respondents commented that competition among staff occasionally got in the way of knowledge sharing, partly because of those compensations. This was found to be an emergent property of the QCN implementation that has negative consequences.

> *“…. remember these health workers in maternity, antenatal and the child clinics all work in shifts. So, I am going to pick nurse A and leave out nurse B in the hope that nurse A will come and tell nurse B what is supposed to be done. They actually don’t even share. The health workers are saying that those who go are getting paid allowances and when they come back, even if they wanted to share, others are really annoyed that they are not selected*.*” [Technical Partner-National level-Uganda Round 1]*

#### ii. Enhancing the culture of documentation and data use

A strong theme to emerge across the four countries was that a critical achievement of the network to enhance health care professional’s documentation culture, aligning with a core network objective to reduce maternal and newborn mortality through use of easily retrievable, local data. As explained by a midwife in Ethiopia:

> *“…. If you don’t record the number of mothers you assisted, you don’t know how many deliveries you have attended. This system has helped me and fellow midwives to improve our documentation culture…” [Health Facility Worker-Local level-Ethiopia Round 1]*

Network countries put renewed emphasis on training for reporting systems as well as learning from identified problems. This was explained to be a major component of capability for QI in Bangladesh where there was a focus on introducing updated data systems and training hospital staff to use them.

> *“The data from different sectors come to the statistician and the statistician updates the database. We couldn’t do this thing before. Now the sisters [nurses] have been motivated to do these after arranging training and doing many things*.*” [Implementing partner-National level-Bangladesh Round 1]*

Despite improvements, actors working at different levels in the system reported that poor documentation practices, poor quality of data collected, lack of data collection tools, lack of knowledge on how to use them, and lack of a culture of data use for decision making continued to be a challenge across the network. In Uganda there were complaints that the ministry of health monitoring tool was not easy to use due to its bulkiness and requiring extra time of providers who are already overburdened, resulting in implementing partners using their own tools: this duplication negatively affected motivation around data use. Numerous respondents in Ethiopia and Malawi reported that their DHIS-2 reports remained incomplete, and mistakes were frequently found despite improvements in report quality. A respondent at one of the health facilities in Malawi elaborated this as follows,

> *“There are data issues, missing data, poor quality data and lack of ownership for the data. People in the facilities should sit down to look at their data… they should ask ‘Is it making sense to us? What are the problems?’ So, there is a lot of work out there to improve*.*” [Government-National level-Malawi Round 2]*

#### iii. The importance of leaders to motivate individuals

Strong leadership was cited as playing an instrumental role in supporting individuals to contribute positively to QCN across the network countries. The active participation of leaders at all levels brought attention to the importance of quality of care, motivating health staff and communities alike.

> *“……About leadership, when the Civil Surgeon puts a focus on, that facility will undoubtedly perform well. The reason for this is that no matter what you want to do, good institutional leadership is critical*.*” [Implementing partner-National level-Bangladesh Round 4]*

However, respondents across the countries indicated that leadership and accountability lacked uniformity between different levels of the network, with different patterns by country. In Ethiopia the network members’ engagement seemed strong at the top and most peripheries; in Uganda QCN seemed more cohesive at the national level compared to the subnational level; in Malawi, network leadership was more robust at the top level. Different levels in Bangladesh showed varied leadership commitment. All countries shared that the continuous reshuffling of leaders negatively affected engagement. Lack of QI knowledge among individuals in leadership positions within health facilities was also cited as a challenge.

> *“Leadership is also an issue in most hard-to-reach facilities. They have people who are just coming from school, they don’t have the experience on management…So somehow, you will find that the leadership to be weak and they don’t even grasp the concept of quality. So, for them to support the QI initiative is a challenge. Once the leadership is weak or not conversant on some of the QI initiatives, you will find that we don’t manage our facilities the way we would want*.*” [Government-National level-Malawi Round 3]*

## 2. Organizational circumstances

Three themes emerged under organisational circumstances [i] inadequate staffing levels; [ii] the need to find strategies for continuously building staff skills; and [iii] the problem of limited physical resources and processes in facilities.

### i. Inadequate staffing levels

Numerous respondents across the four countries said that the inadequate number of healthcare personnel reduced the ability of a facility to engage with QCN effectively. The problem also extended to support staff such as cleaners. This often led to staff performing ‘double duties’, unsanitary conditions, and health care users experiencing long waiting times. For example, findings from an observation in one of the case study sites in Uganda indicated that there was a 1:>50 ratio of provider to patients and, there a 1:12 ratio of providers to beds in the maternity ward. Despite this, some institutions reported success in establishing targets for an adequate number of healthcare personnel by recruiting staff especially in districts.

> *“We have an indicator called ‘approved posts filled with qualified people’. This one has improved really for us. Now we are at 74% …. of course, we cannot avoid the shortages all in all, but I think there is a lot of improvement in the wage bill and recruitment of staff especially in districts*.*” [Government-National level-Uganda Round 2]*

Of concern, nepotism emerged as a challenge mentioned for lack of adequate and skilled professionals in some cases. There were reports of managerial positions and project related work with extra incentives and benefits being monopolized by a small group of people in an institution.

> *“Our current situation is serious. Academic competence is given less attention and people are assigned by race and tribe. They form secret networks and influence the entire institution. Since the leader is not competent enough, he will assign one or two people to support him in every aspect. These people will be assigned to 5-6 programs each. When these people are gone, everything is lost*.*” [Government-Regional level-Ethiopia Round 2]*

### ii. Finding strategies to continuously build staff skills

QCN actors commented on opportunities for building skills of current staff that need either new training or refresher training on a continuous basis. Examples were provided at multiple levels: coaching within supportive supervision, mentoring between facilities, skills building in clusters of facilities, and having visual posters or banners in facilities that show processes. In addition, institutional collaboration with different universities and professional associations was described as a good opportunity for support of staff capacity in facilities in Bangladesh and Uganda. Respondents across the countries agreed on the importance of network activities that brought staff together to support continuous capacity building. A local level respondent from Malawi explained,

> *“We have Continuous Professional Development (CPD) sessions where we invite key players in MNH activities in these 8 facilities where we discuss common conditions that we think need more knowledge and skills so that we will be able to improve the quality of services that we are giving to our patients*.*” [Health facility worker-Local level - Malawi Round 1]*

Building from previously existing initiatives and programs of country health systems were also identified to be important. Participants acknowledged the complementarity of these programs with QCN.

> *“QI initiatives by the MoH are already in the system. We have these… learning districts that report regularly is the new thing the network brought. So, the programs complement one another…. Now hospitals support health centers*.*” [Government-Regional level-Ethiopia Round 1]*

### iii. Limited physical resources and processes in facilities

Lack of physical resources – infrastructure, basic amenities, and equipment - was frequently mentioned to prevent adequate care, or to harm patient satisfaction. Providers reported that this problem had tied their hands to exercise the skills and knowledge they acquired through various trainings.

> *“The implementing partners have done a lot of training on quality of care however on many occasions, health workers are not able to utilise this acquired knowledge due to lack of essential equipment/resources*.*” [Government-National level-Uganda Round 2]*

One respondent from Bangladesh explained how the combination of shortages in staff and resources limited the potential for quality improvement:

> *“So, what is the reason behind this high maternal mortality? The reason is quality has been compromised. You want to increase the delivery rate of the hospitals without the quality improvement? You need the capacity, manpower, system design and different tools. One of my staff reported to me that ‘Sir, I have two death cases of PPH’. I told him to ‘Investigate it*.*’ He came back after 30 minutes. It was found that the waiting time for those two PPH patients was more than 2 hours*.*” [Technical Partner-National level-Bangladesh Round 1]*

Despite the challenge of physical resources, some improvements in processes were noted due to QI interventions, for example maternal and perinatal death, surveillance and response [MPDSR] and death audits in member countries were reported to have been strengthened as a result of QCN.

> *“The audits are conducted more frequently, and the quality of data has improved*.*” [Government-Local-Malawi Round 1]*

## 3. System circumstances

Four themes emerged under system circumstances: [i] the opportunity of policy alignment and partner coordination; [ii] the burden of parallel reporting systems; [iii] paying for quality health care; and finally [iv] managing disruptive events.

### i. Policy alignment and partner coordination

QCN was frequently commented to align well with government policy and rhetoric and to promote partner alignment. QCN purposively identified and engaged NGOs that work on QI areas and quality was where these organizations interface. Respondents considered that government leadership for QCN had been strong in all of the four network countries. It was a slow start in Uganda but later on picked up with the re-structuring of the ministry of health. In Ethiopia, the programme was considered a flagship initiative and there was effective partner coordination. A respondent explained:

> *“The ministry is taking this initiative as a flagship initiative and in fact, other new learning initiatives are replicating this approach. So, this is an overall mobilization from top to bottom level. Most importantly, I believe it is because of the network that the partner organizations are able to implement the national strategy MNH quality of care road map according to the governmental recommendation and in a harmonized approach*.*” [Implementing partner-National level-Ethiopia Round 1]*

By the same token, MNH was a priority area for the Malawian government and the goals of the QCN align with national priorities which facilitated uptake of QCN.

> *“Issues of maternal and newborn health are a priority for Malawi. So, any opportunity that comes, that is geared towards quality services, Malawi is very much interested in that. So, when this network came, it was structured to ministry of health to buy it. Initially it was WHO to talk about the quality of care network. And Malawi got interested and wanted to be one of the countries to get the network*.*” [Government-National level-Malawi Round 2]*

### ii. The burden of parallel reporting systems

Parallel reporting systems were identified as a challenge to continuous, timely and quality data across the countries. One consequence observed in Ethiopia was that implementers were selective in the indicators they engaged with. Indicators related to maternal mortality rate, neonatal mortality rate and stillbirth became their focus areas. Likewise, respondents also mentioned that the indicator “experience of care” was difficult to collect as it required interviews with service users. Similarly, not every public facility in Bangladesh was using the full set of QCN indicators. As an implementing partner respondent from Bangladesh explained, the rollout of quality of care indicators required a high level of complexity:

> *“…But what we could not do still now is bringing the data under national system, neither at DHIS2 nor Director General of Management Information System*.*” [Implementing partner-National-Bangladesh Round 2]*.

Respondents also commented that the parallel reporting system and the documentation process were burdens for health care providers.

> *“This is a very good tool that was developed in partnership with the MoH, partners, even in consultation with the other offices not only in Malawi. And good results came out but in some places it’s too cumbersome for the health unit to complete. But it provides a good baseline and a good tool to monitoring progress*.*” [Technical Partner-National-Malawi Round 3]*

However, regardless of the challenges, some partners had worked successfully with local governments to align the national data systems.

> *“Numerous implementing and monitoring stakeholders, notably UNICEF, Save the Children, and QIS, have worked to include QCN indicators in the formal chain of quality data reporting*.*” [Health facility worker-Local level-Bangladesh Round 2]*

### iii. Paying for quality healthcare

Network countries that made maternal and newborn health a pillar of the health system commented on the importance of financing care. Free maternal and newborn health services, as was the case in Ethiopia, Malawi and Uganda were thought crucial. In the same vein, the result-based financing programme in Uganda was thought to have had a positive impact not only on increased uptake of institutional delivery, but also through improved birth and death registration, motivation of health workers, and enabling facilities to buy supplies.

Nonetheless, all countries reported that direct or indirect health care costs borne by the user were a barrier to achieving network goals, with some costs preventing patients from receiving all of the recommended care. This may include having to pay extra for certain tests, like ultrasound, blood testing, or urinalysis, or having to pay for transportation to a physically distant district hospital.

### iv. Managing disruptive events

All network countries reported the challenge of staying focused on quality improvement in the face of disruptive events. The Covid-19 pandemic had created considerable challenge to health care systems everywhere. Immediate QCN implementation was affected, as were plans for future scale-up. Budgets previously designated for other health care provisions were repurposed to Covid-19 emergency operation centers, according to respondents. Members of the subnational network also mentioned the discrepancy in the frequency and intensity of monitoring and supervision activities.

> *“Of course, at that time because of the fear, things weren’t clear. Even, people also said ‘all the money to COVID’. Some of the facilities decided to take the QM [Quality management] money to COVID. So, there were those situations in which money was channelled to the COVID activities instead of the routine things*.*” [Government-National level-Malawi Round 2]*

But the Covid-19 pandemic was not the only systems challenge reported. The national election of Uganda was said to have temporarily disrupted network operations. And the unstable political situation in Ethiopia was reported to have had an impact on both the site selection and implementation of the project. Five of the nine implementation regions encountered sporadic conflicts during the implementation process. During those occasions, health-care workers were repurposed for emergency tasks, facilities were demolished, and implementers were unable to obtain complete data.

## Discussion

We examined the capacities and functionality of the Quality of Care Network in four network countries, reflecting on challenges and facilitators at three levels: individual, organizational and system. Three core themes emerged that operated across these levels: [i] the need for high quality staff who were trained, retained and continuously motivated; [ii] avoiding parallel data systems and promoting a culture of local data use; and [iii] leadership at multiple levels to motivate individuals but also to drive system policy and coordinate partner actions. In addition, having access to adequate physical resources, financing health care, and managing disruptive events all emerged as key drivers to network functionality.

Individual level contributions in the network activities were found to be the backbone of successful network implementation but it repeatedly emerged that the individual level must have the support of organisations and systems. The baseline skill of available health care workers and their commitment to providing quality services, as reported in other settings, were crucial [13]. As the world’s public health focus has expanded to incorporate quality of care, the scarcity of qualified health care providers to carry out quality improvement operations is a major concern [2, 14].

Beyond the availability of skilled personnel to conduct health care was the need for operations that provided continuous on-the-job training. QCN activities directly contributed to this need through refresher training and learning forum initiatives that brought actors together for shared learning [15]. The attention that the network placed on tracking progress at the facility level also supported ongoing ambitions to improve data recording and data use, although it was not always possible to avoid duplication. These initiatives were seen to fill gaps that country education systems were not able to address through on-the-job training. Lessons drawn from the four countries indicated that the knowledge and skills obtained through continuous learning opportunities also contributed to address the problem of staff retention by reducing attrition of staff and increasing institutional memory.

Failure to develop effective leadership minimizes the chances of a high-quality work environment, which leads to adverse patient outcomes. Transformational and resonant leadership styles are linked to decreased patient mortality, whereas relational and task-oriented leadership styles are linked to improved patient satisfaction [16]. A similar case study which compared rural resource limited facilities of South Africa indicated facilities with authoritarian leadership style had poor maternal outcomes, less staff motivation and poor client satisfaction, though this might not work all the time [17]. Other literature also indicates that a mix of the different leadership styles that consider organizational contexts could bring the desired effects [18]. The importance of good leadership at different levels to improve implementation was clearly seen in the four selected countries, but it was a limitation of the network functioning that leadership and power were not equally distributed at national and sub-national levels. Beyond direct implementation, political leadership, and commitment to smooth and successful implementation of programs was also key. Partner and government alignment and coalition were crucial to the functioning of QCN in all the four countries [19]. Adequate financing of essential health services is crucial [20]. A bottleneck analysis applied in 12 countries in Africa and Asia indicated that health care financing and health workforce were the top two bottlenecks for scale-up of maternal-newborn intervention packages [21]. Furthermore, in LMICs, infrastructure and basic amenities remain the most significant contextual obstacles to successfully utilizing and benefiting from interventions and similar networks [22]. These issues resonated through our study, showing that, despite some characteristics of the network helping to problem solve, trying to improve quality of care in the absence of an adequate health workforce, infrastructure, country-led finance, or distributed leadership was challenging.

In LMICs the foundation of a health system can easily be shaken because of the little reserve capacity to respond to a shock. The Covid 19 pandemic was a perfect example of this. While contextual adaptation and accommodation of interventions in an existing environment could be a foundation for a sustainable and resilient system, repurposing of health professionals and health facilities during the pandemic has clearly negatively affected maternal and newborn care [23]. Likewise, contextual factors like political instability, as well as natural calamities such as droughts and floods, may impair on-going health efforts by disrupting usual service delivery [24]. In the case of QCN, such challenges directly linked to limiting the possibility of conducting learning sessions, reducing the pool of available staff, restricting transportation and supply chains, and reducing service provision.

## Strengths and limitations of this study

Research methods that were harmonised between time and place across four diverse countries represented a unique opportunity for learning. However, implementation of the research methods was driven by country specific local realities meaning that the learning was not uniform between settings. In addition, the framework used for this paper has a limitation in not focusing on what many have called the invisible software of health systems and the network such as intra and inter-professional relationships, local ‘practical norms’ and other aspects of local facility culture or local cultural issues. Nonetheless, the consistency of themes emerging across these four different contexts suggested a high level of external validity and potential of extrapolation to inform future health networks.

## Conclusion and recommendation

Implementing multicounty networks in different settings necessitates inputs and coordination at multiple health system levels. To achieve the network’s objectives, it is necessary to have an adequate amount of health system financing, strong political leadership, and coordination, as well as dedication to streamlined data systems, which requires organizational structures filled with motivated and well trained individuals who benefit from local leaders and have a positive data use culture. Within countries, the intended network goal to reduce maternal and newborn mortality had the best opportunity to be realized through coordinated efforts of subnational and national stakeholders. In the selected four network countries, the characteristics of QCN helped to address issues related to staff, leadership and data use. Capacity challenges encountered at individual, organisational and system levels hampered the initiative’s ability to succeed as intended. Nonetheless, sufficient gains have been made to warrant investing the time, effort and resources needed to sustain the Network rather than introducing other new programmes.

## Data Availability

All data produced in the present work are contained in the manuscript

## Acknowledgment

We would like to forward our gratitude to the national and local level network implementation stakeholders and members including the QCN technical working group members in Bangladesh, Ethiopia, Malawi, and Uganda for their relentless support during the data collection process. We thank all respondents and stakeholders for their time and contributions toward making this work possible. The QCN Evaluation Group is: Mithun Sarker, Abdul Kuddus, Kishwar Azad [BADAS-PCP Bangladesh], Albert Dube, Gladson Monjeza, Rachel Magaleta, Zabvuta Moffolo, Charles Makwenda [Parent and Child Health Initiative, Malawi], Mary Kinney, Fidele Mukinda [independent researchers, South Africa], Yusra Shawar, Will Payne, Jeremy Shiffman [Johns Hopkins University, USA], Agnes Kyamulabi, Hilda Namakula, [Makerere University, Uganda] Asebe Amenu, Theodros Getachew, Geremew Gonfa [Ethiopia Public Health Institute, Ethiopia].

## Supporting information

**S1 Text. PLOS Global Public Health QCN Evaluation Collection 2-page summary**.

**S2 Text. PLOS Global Public Health QCN papers common methods section**.

