## Supplementary material for "Individual, organisational and system capacities, and the functioning of a multi-country implementation-focused network for maternal, newborn and child health: Bangladesh, Ethiopia, Malawi, and Uganda": S2

#### QCN EVALUATION METHODS SUPPLEMENT

This document provides details of our methods across the whole of our QCN evaluation project and is therefore a common resource for all 9 papers in the PLOS Global Public Health collection [ref PLOSGPH collection website]. Section 1 provides an overview of our research questions, objectives, and the evaluation outcomes we are focusing on across the whole project. Section 2 describes each study setting. Section 3 describes our data collection methods. We used interviews, document review, observations, and surveys at local and national levels within Bangladesh, Ethiopia, Malawi, and Uganda (our case study countries) and at global level, with participants and events purposefully selected to best meet our objectives (Figure 1). Section 4 describes our analytical methods, which draws on our theoretical framework described in Section 5. Section 6 focuses on ethics and ethical approvals.

#### 1. RESEARCH QUESTIONS, OBJECTIVES & EVALUATION OUTCOMES

***Research questions:*** We examined how QCN was constructed, its operations and their effects, focusing on network actors (individuals and organizations). We examined what aspects of the QCN work best and how it influences global, national, and local levels by tackling three research questions (also see Figure 1):

1. **Global level**: What attributes of this multi-country network and its operational strategy and performance affect the engagement of network actors at global and national levels and their adoption of a shared agenda and goals to improve maternal and newborn health services?
2. **National level**: What shapes the relationship between country teams and the global network leadership and how does this influence ownership of the policy and management work that is required to set national aims and improve services, and which characteristics of the health system context appear to influence this?
3. **Local level**: What specific form does national QCN activity take and how does this influence which specific interventions are delivered, which of these are felt to be successful by local actors and which lead to measurable changes in processes and outcomes?

We attempted to answer our three research questions via a multi-disciplinary mixed methods programme of work aiming to achieve the six objectives detailed in Figure 1 (see section 4 below for theory) focusing on the QCN global level, and the national and local programmes in our case study countries Bangladesh, Ethiopia, Malawi, and Uganda. We sought to produce generalisable theory relating to QCN operations at global, national and local level.[1]

**Figure 1** Environment, Structure, Process, Outcome model of network and proposed evaluation objectives [and methods]

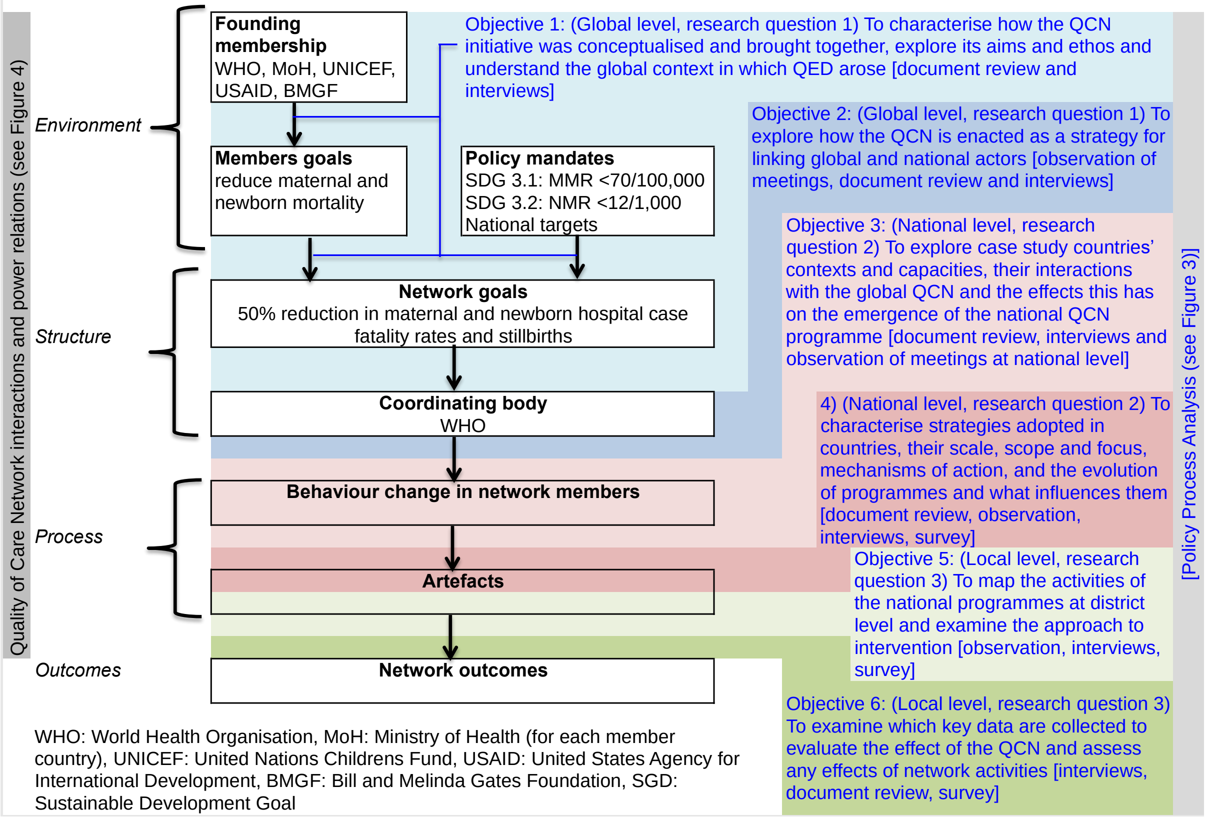

***Evaluation of outcomes:*** We aimed to investigate how the network functions – i.e., whether it was 'working', at global level, at national level in our selected case study countries, and at local level in selected health facilities in our case study countries. The main outcomes evaluated in our research were:

1. At global and national levels: adoption of a shared agenda and goals to improve maternal and newborn health services (**network emergence**)
2. At national level: ownership of the policy and management work of the network (**network legitimacy**)
3. At local level: delivery of locally valued interventions (**network effectiveness**)

These correspond to our three research questions stated on page 1. Our research is predominantly focused on why and how actors work to achieve desired outcomes and how this explains the degree to which they are achieved, using the methodology described below. Whether the network is aligned with national priorities or will have good policy or service-related outcomes given opportunity costs, the potential for creation of boundaries around certain actors, exclusion of other relevant stakeholders or unintended effects, are all open questions that we explored. The answers to these questions are needed for decision-making on future global networks. Whether the network reduces maternal, neonatal and stillbirth case fatality rates in participating health facilities is a related question and beyond the scope of our work unfortunately.

#### 2. STUDY SETTINGS

The situation in each of our case study countries is different with respect to political engagement, and on-going and planned activities related to maternal, newborn and child health that could be leveraged or that present barriers to the successful emergence, legitimacy, and effectiveness of QCN in the country. These situation in each country prior to the start of our work in 2019 is described in brief below to add context to our work and explain the relevance of our case studies. Further detail, including a description of the evolving context in each country is included in our country context supplement.

**Bangladesh**: The Government of Bangladesh (GoB) is committed to achieving SDG targets and applies a sector-wide approach under the national Health, Population, and Nutrition Sector Plan 2017–2022 to address the needs and gaps for accelerated progress in maternal, newborn and child health (MNCH).[2] The current operational plan includes activities to strengthen efforts to: make home deliveries safe; provide *24/7* emergency obstetric and newborn care services at the upazila (sub-district) level in phases; establish a functional referral system from community to facility level; and, increase access to and utilization of evidence-based priority newborn and child health services.[2] The major challenges identified are: to ensure deliveries by skilled birth attendants; to reduce unnecessary caesarean section deliveries by private health facilitates; to reduce significant inequalities of service utilization across geographical regions and between different wealth quintiles; to make ready union level health facilities to provide normal delivery care services; and, to reduce neonatal deaths.[2] The union level facilities offer an actionable opportunity to strengthen the provision of life-saving care to mothers and newborn during and at the time of birth. To ensure the quality of MNCH services, a functional leadership structure for quality improvement has been established and a national quality of care strategy for the health sector including for maternal and newborn health services has been developed. Quality of care committees for district health management teams are currently being established.[3]

**Malawi:** Malawi was one of the few countries to achieve its Millennium Development Goal target for child health. It adopted Every Newborn Action Plan in 2015 and is currently reviewing its Reproductive, Maternal, Neonatal, Child and Adolescent Health (RMNCAH) strategy. Institutional deliveries have increased significantly in the past few years and have outpaced increases in skilled human (and material) resource availability. Quality of care for many mothers and newborns remains poor as a result. The Government is cognizant of this and has recently established the Quality Management Directorate (QMD) within the Ministry of Health. QMD aims to contribute to improved health and client satisfaction via provision of quality health services in Malawi and has three divisions: standards and norms (development and promotion of quality standards and guidelines), quality improvement (identifying and supporting quality initiatives) and monitoring and evaluation (assessing quality initiatives, and provision of supportive supervision). QMD is involved in QCN. The Government's commitment to quality is also reflected in the Health Sector Strategic Plan (HSSP-II). Under the leadership of QMD, Malawi has developed its National Quality Policy and Strategy (NQPS). There is also strong donor commitment to support the quality agenda in the health system, specifically for RMNCAH. Improving quality of care however requires a culture shift, which can be slow. Greater investments of time and resources are required for integrating quality initiatives into the health system in Malawi and fostering a culture of learning.

**Uganda:** While the rate of facility deliveries, skilled deliveries, and utilization of health facilities for curative care has increased in recent years, there are gaps in the quality of care provided. Quality of care is central to the global agenda of ensuring health for all at all ages (SDG 3), a goal shared by the government of Uganda. The adoption of various components of quality in healthcare dates back to 1994 in Uganda; it was the first country to implement quality assurance on a large scale in Africa.[4] Several quality management interventions have since followed including the Yellow Star program, professionals' registration, licensing and accreditation, infection control and prevention. In the recent past, the national standards on maternal and newborn health (MNH) quality of care (QoC) as well as the health sector quality improvement (QI) framework and health sector strategic plan 2015/16–2019/20 have been developed; these may result in the institutionalisation of QoC initiatives nationally. The MoH has also began to implement national QI interventions including QI coaching, clinical mentorship and audit and feedback in select districts.[3] These have been supplemented by externally funded projects that have taken on some of these interventions across different regions in the country.[5, 6] Uganda also has plans to improve data systems, create learning networks and systems and performance-based-financing to facilitate QoC at different health system levels. Formative work is underway within selected districts. Progress will depend on how effectively system bottlenecks and other key issues are conceptualised and addressed. Current challenges include inadequate execution of QoC interventions especially perinatal death audits and mentoring nationally; a lack of harmonisation between MoH and other players; and a lack of standardised reporting and evaluation mechanisms. Community preferences and value systems have also yet to be incorporated into current narrow standards-driven approaches to improving the quality of health services in Uganda.[7]

**Ethiopia:** Maternal and new-born mortality remain high in low-resource settings, including Ethiopia. With increasing rates of births in hospitals in Ethiopia, there is a need for health system interventions that optimise quality of care so that further reductions in mortality can be achieved despite resource constraints. The country's health sector transformation plan also set goals to improve quality of health care and utilization of essential health services (12).
Despite the gains that Ethiopia's health system has achieved in a short period of time, more can be done to improve maternal and child health outcomes. Thus, HSTP stipulates that the government of Ethiopia must continue to prioritize the improvement of reproductive, maternal, newborn, child, and adolescent health services, as indicated in the sustainable development goals (SDGs), Ethiopia will intensify RMNCAH interventions to end preventable maternal and child deaths by 2030 (12).

As part of our case studies, we have also focused our analysis on the global level and how it operates and interacts with the national level in our case studies. At the global level, the QCN is steered by the WHO's department of Maternal, Newborn, Child, Adolescent Health, and Ageing, acting as the global convenor, coordinator, and Secretariat for the network. The QCN secretariat is further divided into three working groups: Implementation & Learning, Monitoring & Evaluation, and QED Advisory Group. Whilst the WHO lead the QCN Secretariat, several global partners contribute to global activities, funded by the Bill and Melinda Gates Foundation (BMGF). For example, the working groups include actors such as UNICEF, URC ASSIST, IHI, Save the Children, UNFPA, MSCP, and Jhpiego, that bring their own resources in terms of knowledge and time. UNICEF and USAID are important global actors involved in the implementation of QCN activities in-country as UNICEF received funding from BMGF for implementation in-country and USAID acts as an investor through their different projects and partners in several QCN network countries.

#### 3. DATA COLLECTION METHODS

##### 3.1 Interviews

We conducted semi-structured interviews[8] with national level (objectives 3, 4) and local level (objectives 5, 6; Figure 1) network members and key stakeholders in Bangladesh, Ethiopia, Malawi and Uganda. We sought to pay particular attention to the perspectives and goals of those carrying out the work of the network.[9, 10] In each country we conducted several iterative rounds of interviews, at least six months apart, to capture changes in how the network was operating and views pertaining to network activities as well as follow-up on emerging findings from the previous round. The topic guides used in interviews were adapted to each level of stakeholders and to each country's specific context, and further iterated through the rounds (examples of first round topic guides for national and local level interviews are provided as Appendices 1 and 2 at the end of this document). Topic guides were also translated in local languages as necessary. The number of rounds in each case study country and the number of interviews with national and local level stakeholders in each round are provided in Table 1. Interviews were conducted by members of the QCN Evaluation Team trained in qualitative data collection methods who were also familiar with the local contexts and languages.

**Table 1: QCN Interviews completed (including follow-up interviews)**

| Case-study Country | Data collection Round (dates) | National Level interviews (n) | Local Level interviews (n) |
|---|---|---|---|
| Bangladesh | 1 (October-2019 – March-2020) | 13 | 07 |
| | 2 (Oct-2020 – Jan-2021) | 14 | 11 |
| | 3 (May-2021 – Sep-2021) | 10 | 12 |
| | 4 (Jan-2022 – March-2022) | 08 | 00 |
| Ethiopia | 1 (Dec-2020 – Mar-2021) | 08 | 11 |
| | 2 (Sep-2021 – Dec-2021) | 10 | 11 |
| Malawi | 1 (Oct-2019 – March-2020) | 07 | 12 |
| | 2 (Nov-2020 – Jan-2021) | 10 | 07 |
| | 3 (Aug-2021 – Nov-2021) | 09 | 07 |
| | 4 (Mar-2022 – May-2022) | 04 | 03 |
| Uganda | 1 (Nov-2020 – Mar-2021) | 07 | 13 |
| | 2 (June-2021 – Sept-2021) | 12 | 08 |
| | 3 (Feb-2022 – Mar-2022) | 10 | 05 |

We also conducted two rounds of interviews with global level (objectives 1, 2) network members and key stakeholders. The first round was conducted in March 2019 at the QCN global meeting in Addis Ababa, Ethiopia and seven people were interviewed including representatives from WHO, the Bill and Melinda Gates Foundation, USAID and IHI. The second round of global level interviews was conducted towards the end of our QCN evaluation project between November 2021 and January 2022, and 14 people were interviewed (see Appendix 3 for topic guide).

To test our ideas and inform development of our proposal for this work we also conducted preliminary interviews with 18 MoH and global stakeholders from a number of the QCN countries at the second QCN meeting in Dar es Salaam in December 2017.

##### 3.2 Document review

We reviewed accessible published and unpublished documents and communications relating to the QCN at global level (objectives 1, 2) and at national (objectives 3, 4) and sub-national (objective 6; Figure 1) levels in the case study countries. These included strategy and management documents, operational plans, directives, formal minutes, and reports (Table 2). We were able to access unpublished documents via WHO and Ministry of Health QCN contacts. We analysed the content of the documents using the same coding framework in NVivo as for our interview data, as described in our analysis section below.

**Table 2: QCN document reviews completed**

| QCN Evaluation Level | Document Type | Number of documents reviewed |
|---|---|---|
| Global | Strategy document | 5 |
| | Operational plan | 5 |
| | Report | 5 |
| | Minutes | 0 |
| National - Bangladesh | Strategy document | 3 |
| | Operational plan | 1 |
| | Report | 5 |
| | Minutes | 35 (presentation 8) |
| National - Ethiopia | Strategy document | 4 |
| | Operational plan | 6 |
| | Report | 7 |
| | Minutes | More than 20 |
| National - Malawi | Strategy document | 1 |
| | Operational plan | 2 (presentation 1) |
| | Report | 9 (presentation 2) |
| | Minutes | 6 |
| National - Uganda | Strategy document | 3 |
| | Operational plan | 2 |
| | Report | 9 |
| | Minutes | 0 |

##### 3.3 Observations:

We conducted non-participant observations[8] of multi-country meetings (objective 2) and key national-level (objective 3) and district level (objective 5) meetings in case-study countries. Activities at district level were also observed via visits to two better and two least performing QCN hospitals in each case study country (objectives 4, 5) in several iterative rounds (Table 3). Best and least performing facilities were selected based on maternal and newborn health outcomes and other quality of care data (e.g., those used in national schemes) relevant for each country. We used templates (e.g., Appendix 4) to capture key processes relevant to the focus of the network in each country during observations, as well as unstructured notes.

The observations in health facilities were used: i) to explore whether what people say is being done is what is being done, ii) whether those at the frontline feel anything has changed over the period of QCN operation, iii) to explore why activities are done (or not done), and iv) to explore any effects and the veracity of monitoring data (objectives 4-6).

Our sampling strategy for these observations was therefore based on ensuring a diverse sample of facilities and health worker cadres in each country rather than ensuring statistical representativeness. To ensure our observations were informative they were conducted by trained and experienced researchers familiar with the local setting (culture, language, context), and recorded in detailed field notes and the focus of observation was sharpened over time enabled by iterative rounds of data analysis and reflection.

**Table 3: QCN observations completed**

| Location | Observation Type | Rounds of observations (total days) |
|---|---|---|
| Bangladesh | Best performing facilities | 3 rounds (6 days) |
| | Least performing facilities | 3 rounds (6 days) |
| | QCN national meeting | 1 round (3 meetings) |
| | QCN local (district) meeting | 0 |
| Ethiopia | Best performing facilities | 2 rounds (7 days) |
| | Least performing facilities | 2 rounds (only visited one of the facilities) |
| | QCN national meeting | 1 |
| | QCN local (district) meeting | 0 |
| Malawi | Best performing hospitals | 3 round (6 days) |
| | Least performing hospitals | 3 rounds (6 days) |
| | QCN national meeting | |
| | QCN local (district) meeting | |
| Uganda | Best performing hospital | 3 rounds (6 days) |
| | Least performing hospital | 3 rounds (6 days) |
| | QCN national meeting | 1 meeting |
| | QCN local (district) meeting | |
| Global level | QCN global meetings | 1 round (3 days) |
| | QCN global webinars | 6 webinars |

##### 3.4 QCN Survey:

We adapted a psychometrically validated tool (5 domains, 40 indicators) developed for evaluating clinical networks[11] to evaluate the network at national (objective 4) and local (objectives 5, 6) levels in Bangladesh, Ethiopia, Malawi and Uganda (example shown in Appendix 5, the survey was adapted for use in each country). We conducted several rounds of the survey in each country (Table 4) and in each round a wide variety of network member cadres (clinicians, managers, advisors) were surveyed. The survey was more widely completed by a far higher proportion of the people involved in the network than those specifically targeted by our interviews and observations. Surveys were administered using paper or online via the UCL Opinio platform following email invitation.

We also conducted a separate stakeholder network survey – the details of this can be found in the paper in this collection dedicated to this by Mukinda and colleagues.

**Table 4: QCN surveys completed**

| Case-study Country | Survey Round (dates) | Surveys completed (n) |
|---|---|---|

| | | |
|---|---|---|
| Bangladesh | 1 (October 2019 – December 2019) | 133 |
| | 2 (December 2020 – January 2021) | 163 |
| | 3 (June 2021 – September 2021 - longer period due to Covid restrictions) | 151 |
| Ethiopia | 1 (Jan 2021 – Feb-2021) | 174 |
| | 2 (Nov 2o21 – Dec 2021) | 190 |
| | | Feb-March 2022 (45 SNA survey) |
| Malawi | 1 (Nov-2019 – Jan-2020) | 119 |
| | 2 (Oct-2020 – Jan-2021) | 191 |
| | 3 (April-2021 – June-2021) | 135 |
| Uganda | 1 (Nov-2020 – Dec-2020) | 139 |
| | 2 (June-2021 – Aug-2021) | 130 |

##### 3.5 QCN monitoring data review

We critically appraised country-level reporting on processes and outcomes[12] and whether and how these data are used for decision-making at global, national and local levels. This data was only made available toward the end of our work and was not detailed enough for us to adequately assess the plausibility[13] of network effects on health outcomes in relation to the evolution of network activities in country as we had originally planned to.

#### 4. ANALYSIS

In this section we first describe our analysis of the interviews, observations and documents using a common coding frame. Then we describe our analysis of the survey data. Finally, we describe our policy process analysis. The following section details the theories all of these analyses draw on. Our analyses were iterative, treating the global level and each country as a specific case, to explore emerging findings while drawing on initial and framing theories[1] (see section 5), i.e. using both inductive and deductive approaches.[8] We answered all three of our research questions related to QCN emergence, legitimacy and effectiveness (see section 1) using these analyses and an overall synthesis of all case study data.

##### 4.1 Analysis of Interview, Observation and Document review data

We used a common coding framework developed from the underlying theories pertinent to our work (see section 5) to code the qualitative data we obtained from the interviews, observations, and document reviews we conducted. All data was coded in NVivo 12, drawing on our initial theories in both an inductive and deductive way. Our codebook contained 'theory' codes related to all the underlying theories described in section 5; each theory was outlined using codes and sub-codes that broke down the different components of the theory. The codebook was further supplemented by 'case study' codes to distinguish data specifically relevant to each case study. The codebook was initially piloted on a set of 14 interviews, conducted with national QCN actors from eight network countries during the international meeting in Lilongwe, Malawi in 2017 that launched the network, as part of our baseline study. The codebook was then tested on the first round of global interviews collected in March 2019 (section 3.1). The initial piloting and testing of the codebook was conducted by five senior co-investigators, whereby two interviews were separately coded by all coders with results later compared and discussed within the team. The remaining of the interviews in the pilot phase were coded by at least two coders with results and discrepancies discussed as a team to refine the codebook as needed.

Our codebook was then shared with the remainder of the research team. It provided researchers with the list of codes for each theory and case study, with a detailed description of each theoretical unit and each code, as well as reference papers for further reading. All researchers involved in the data collection and/or data analysis received a series of trainings on the NVivo project and codebook by two senior co-investigators involved in developing the coding framework. Each new coder started with a small set of interviews that were also coded by another researcher, with results discussed among the wider team to ensure standardisation of the coding. Over two years, many of our research team were involved in the coding of the qualitative data in the following way:

- 7 researchers coded the data from the Bangladesh case study (including 3 actively involved in local data collection),
- 2 researchers coded the data from Ethiopia that were both actively involved in local data collection,
- 8 researchers coded the data from Malawi (including 5 actively involved in local data collection),
- 6 researchers coded the data from Uganda (including 4 actively involved in local data collection),
- 3 researchers coded the global level data (including 1 actively involved in data collection).

Regular team meetings and rechecks took place during each round of coding in order to ensure inter-coder consistency and that coders remained close to the theories underpinning the codebook. Team meetings were also an opportunity for coders to put forward new codes they deemed relevant to refine the 'theory' codes or to address gaps in 'case study' codes. Agreed new codes were then added to the global team codebook and NVivo project, with a detailed description, to be used in the next round of data analysis.

Following coding of each data round, coders wrote up the results of their analysis in a living document (one for each case study), accessible to the whole research team. Living documents were updated and refined following each round of data analysis and further finalised after the last round of analysis. Those living documents served as a basis for the writing-up of the papers in our collection. Three papers in our collection (Akter et al., Tesfa et al. and Lemma et al.) additionally conducted thematic analyses for the purpose of answering in more depth adjacent research questions. Those analyses are described in the methods section of each paper.

##### 4.2 Survey analysis

Survey questions were framed both positively and negatively so that framing does not bias responses and people need to read all questions rather than just ticking the same 'agree' or 'disagree' response down a column covering several differently framed questions in the same section.

We report descriptive statistics of respondents and the percentages of respondents giving particular responses to each question. We calculated summary scores for each question (indicator) by scoring strongly positive responses as 2 ('strongly agree' to something positive about the network or 'strongly disagree' to something negative), positive responses as 1 ('agree' to something positive about the network or 'disagree' to something negative), neutral responses as 0 ('neither agree or disagree' or 'don't know'), negative responses as -1 ('disagree' to something positive about the network or 'agree' to something negative), and strongly negative responses as -2 ('strongly disagree' to something positive about the network or 'strongly agree' to something negative); and averaging the scores given for the question for all responses. We used these summary scores to calculate domain scores as the sum of the scores for all of the indicators in each domain[11](see Table 5 in the results for how each question maps to each domain). Domain scores were then converted to a percentage of the maximum score possible on a scale from the minimum possible score (e.g.-14 for Domain 6) to the maximum possible score (14 for Domain 6). We also calculate percentage scores for each indicator (question) using this method.

We ran linear regressions of each domain score to see if they varied by respondent characteristics (sex, cadre, place of work). We ran both unadjusted models for each respondent characteristic for each domain score, and adjusted models with combinations of respondent characteristics for each domain score.

##### 4.3 Policy process analysis

We investigated how policies and programmes related to the QCN were formed, implemented, and assessed via examining the interactions between relevant actors and bodies within each country and externally during the agenda-setting, policy formulation, decision-making, implementation and evaluation stages of the policy process, as explained in section 5 below. We investigated the relationships and interactions in terms of policy capacity, power, identity, norms, and related enablers and constraints (see section 5) using a range of qualitative methods including (see above): interviews with key actors and stakeholders, analysis of key documents from involved actors (memoranda of understanding, meeting minutes) and observations of network meetings. We employed a case study process-tracing methodology[14], drawing on and triangulating across multiple data sources to uncover cause-effect mechanisms. We developed our analysis using data from the global level and four national level cases: Bangladesh, Ethiopia, Malawi and Uganda.

#### 5. THEORETICAL FRAMEWORK

Inter-organisational networks, defined here as "*groups of three or more legally autonomous organisations that work together to achieve not only their own goals but also a collective goal*"[15], can be useful to solve *'wicked'* problems in many health systems.[16, 17] They rely on implementation of a shared programme theory[16] – in this case the ToC, developed by the founding partners of the QCN (Figure 2).[12] Notably this ToC does not explicitly include the role of the network, or an appreciation of its capacities (see below). Sheaff and Schofield[16] propose that networks can be conceptualised using a modified version of the Donabedian structure, process, outcome model of quality of care[18], that also accounts for the environment the network was founded in and its members' goals and relevant policies[16]. We adopt this approach and present a simplified example of how it helps structure exploration of the work of the QCN and the attributes we plan to evaluate in our research (see section 1 including objectives in Figure 1).

**Figure 2: WHO-developed Theory of Change for Quality of Care Network**

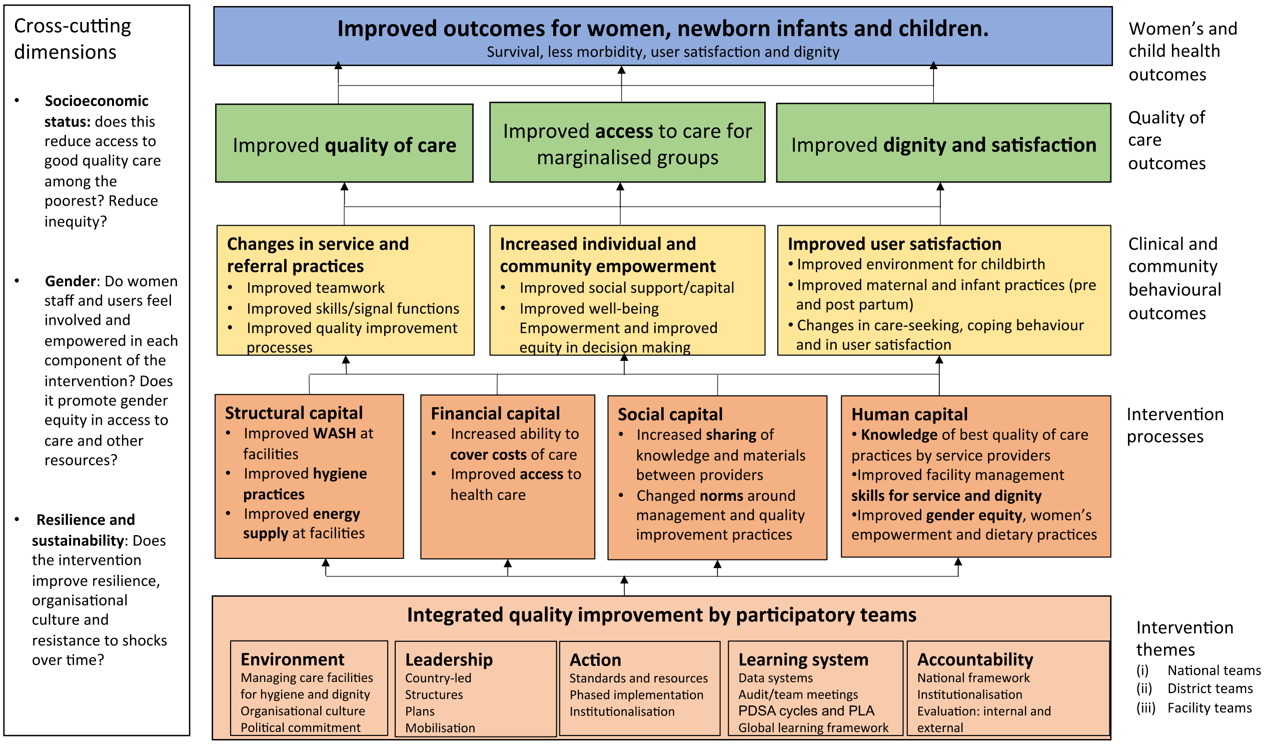

We propose that the QCN can be conceptualised as a 'managed network' [16], where WHO are the lead organisation[15] providing guidance on how to improve the technical quality of care, while also advising on monitoring and evaluation, and organising formal multi-country, multi-stakeholder network engagement. Other partners include funders and external technical agencies supporting quality and systems improvement. The QCN differs from many prior global health networks, which have sought to draw donors' and global actors' attention to specific health challenges,[19] in that its purpose is to *operationalize* quality improvement within countries to reduce mortality (Figure 2).

To address research questions 1 and 2 (section 1), we build on a conceptual framework characterising factors that shape the emergence and effectiveness of global health networks, developed by Shiffman and colleagues.[20] This framework considers "*(1) features of the networks and actors that comprise them, including leadership, governance arrangements, network composition and framing strategies; (2) conditions in the global policy environment, including potential allies and opponents, funding availability and global expectations concerning which issues should be prioritized; (3) and characteristics of the issue, including severity, tractability and affected groups*"[20]. We seek to advance this framework for use with such operationally focused networks as the QCN,[21], extending it to capture dynamics pertaining to implementation by linking it with theories on clinical network functioning and with reference to determinants of success of collaborative approaches to quality improvement.[22-25] For question 3, we were guided by the QCN ToC[12] (Figure 2) and its interaction with the environment, structure, process and outcomes links of the QCN (Figure 1).

We will use recent theoretical advances combining policy-cycle, multiple streams, and advocacy coalition frameworks to understand the policy process[26] to guide our analysis of the agenda setting, policy formulation, decision-making, policy implementation and policy evaluation aspects of the work. This combined theoretical framework for policy-making is useful as it encapsulates the interplay of the various actors, coalitions, influences, and influencers involved as well as the stages of the policy process and critical junctures between them in terms of windows of opportunities when different streams related to problems, politics, potential policy solutions, –and later– the policy-making process and programme implementation coalesce. Figure 3 shows these streams, critical junctures, and the combined policy-making framework in terms of our research foci. It is interesting to note that the authors of the combined policy analysis framework paper conclude by advocating for a cross-case analysis of policy processes in two countries much like we have conducted for four countries as a means to advance theory further[26] – we hope to achieve this.

We are also cognisant of the analytical, operational and political competencies and capabilities required at individual, organisational and system level for ‘successful’ policy-making[27], and therefore also consider these, and their determinants[27] when considering our research questions and objectives. Importantly here we consider the policy capacity of WHO and other multilateral actors as well as the country governments and MoH, and the imbalances between them that may explain, to some extent, the power relationships between them and greater or lesser influence of each on agenda setting and policy formulation in particular (see below and Figure 4). We also draw on capability, opportunity, motivation behaviour change theory (COM-B)[28] here to look at the role of opportunities and motivating factors as well capacities, in determining the manifestation of QCN, it’s activities and outcomes, specifically at the local level (depicted in Figure 5, see below).

Our theory-based evaluation is framed within over-arching theories surrounding the interplay of structure, agency and power[29] (and how this depends on the policy capacities outlined above), diffusion of innovation[30] (information transmission networks with individuals as nodes[31], potentially important for success of quality improvement initiatives[22, 24]), organisational networks with organisations as nodes[31, 32] and transition from hierarchical to network organisation.[17] In Figure 4 we outline potential interactions between different actors and bodies involved, and external to, QCN and how power may be exercised between them, depending on capacity, identity, norms and related barriers and enablers. We explore these relationships in our research

Finally, Figure 5 shows how the different lenses on our work (Figures 1-4) fit together in a simple logic model. This provides an overview of our theory-based evaluation of the QCN. We believe our research is novel as it attempts to span examination of the policy process, including implementation, at the global-national interface with ‘operational’ theories of implementation through organisations and behaviour change theories to ultimately impact frontline workers and users of the health system. These theories follow the anticipated temporal sequence of policy to practice but accept that these transitions are messy; that practice can influence policy and that policy intentions can be blocked. The longitudinal, iterative, and prospective nature of our study has allowed us to tease out some of these dynamics.

**Figure 3 Policy Process Framework linked to wider theories of power and networks**

| Policy Stage | Research question | Policy Stream | Critical junctures for the Network |
|---|---|---|---|
| Agenda-Setting | 1 | Policy solution; Problem; Politics | A. Agenda set: network goals agreed |
| Policy Formulation | 2 | Process | B. Country network policies set |
| Decision-Making | 2 | | C. Country network aims and strategy for achieving them agreed |
| Policy Implementation | 3 | Program | D. Network activities implemented |
| Policy Evaluation | 3 | | E. Network activties evaluated |

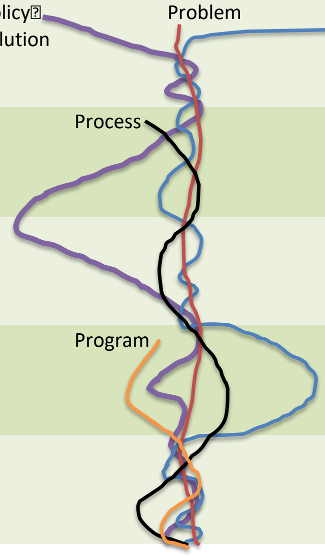

Figure adapted from Howlett et al 2016 [ref 43]

**Figure 4: Quality of Care Network Interactions and Power Relations (to investigate)**

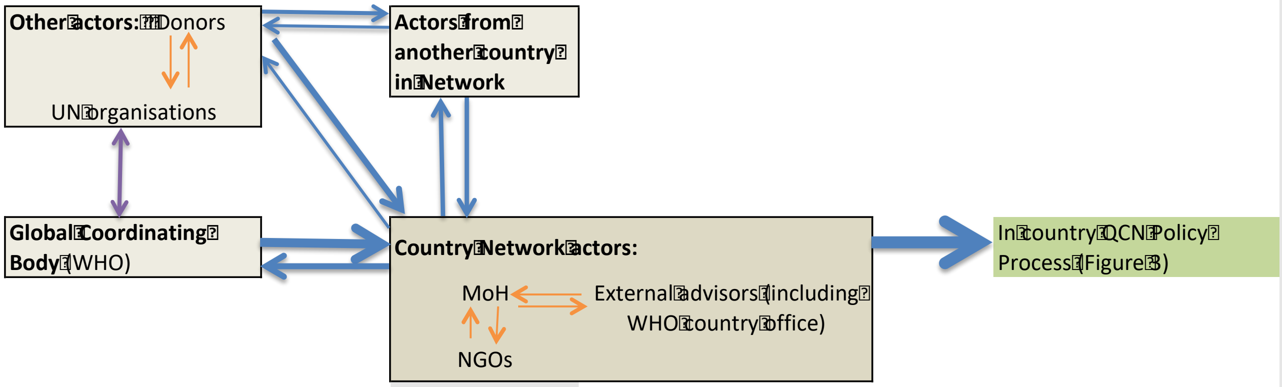

***Extent of influence of each body and actor (width of arrow) will depend on:***

- Capacity [see Wu et al, ref 44] e.g. Human resources available in country to draft policy and programmes, organise activties, analyse and evaluate processes and outcomes
- Alignment of goals between actors
- Organisational culture and wider cultures
- Leverage via other agreements and influences
- Political stability

**Figure 5: Simple logic model outlining how the different lenses and components of our planned research fit together**

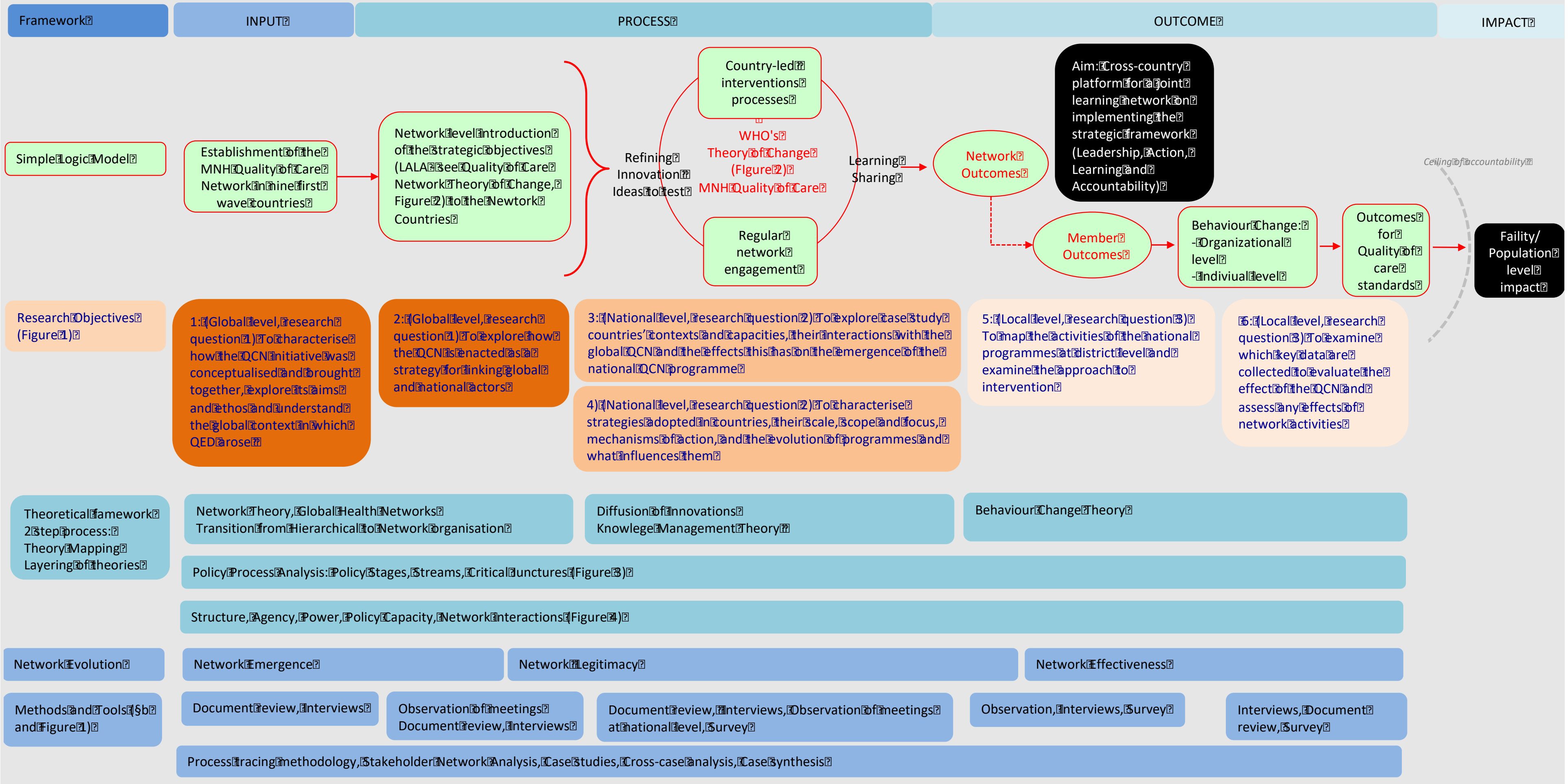

#### 6. ETHICS

All interviews, observations and surveys were conducted after obtaining informed consent, including separate consent for tape recording. Patients' privacy was respected during hospital observations. All data is confidential and anonymised. Ethical approval was obtained from the Research Ethics Committee at University College London (3433/003), National Health Sciences Research Committee in Malawi (Protocol number: 19/03/2264), Institutional Review Board in Bangladesh (BADAS-ERC/EC/19/00274), Ethiopian Public Health Institute in Ethiopia (EPHI-IRB-240-2020) and Uganda (Makerere University School of Public Health Higher Degrees Research Ethics Committee, Ref: 869).

#### Appendix 1: First round national level interview topic guide

**Interview topic guide for Quality of Care Network National level stakeholders**

*Information to read to interviewees first:* We are undertaking a research project on the quality of care network and how it operates. We would like to get the opinion of key stakeholders like yourself on how the network is doing. With your permission, we would like to ask you some questions on the work of the network at global level and in your country, and your personal role in the network and that of your team. This research project is funded by the UK Medical Research Council Health Systems Research Initiative and is being undertaken by Parent and Child Health Initiative in Malawi, Bangladesh Diabetic Association Perinatal Care Project in Bangladesh, Makerere University in Uganda, University College London, Oxford University and Johns Hopkins University, who are evaluating the work of the network as a whole. There are no right or wrong answers or any consequences (positive or negative) of taking part in this interview, and we would really like you to feel free to express your opinions without worrying about what they are. All information will be used to improve the work of the network and future networks. You are free to refuse to take part in this interview and to stop the interview at any time. With your permission we would like to record this interview so we can fully consider all your responses. The recording and any notes I take will be kept secure and you will remain anonymous in any report stemming from this work.

Permission to be interviewed: signature: ___________________________________

Permission for interview to be recorded, signature: ___________________________________

Date: _________________________

Which country are you representing? What is your position?

**The network and your country**

1. Tell me what you understand about how your country got involved in this network? Who seemed to really be leading efforts to join the network (probes: was it government, WHO, UNICEF, other organisations or individuals?)
2. Who is part of the network now in your country? What roles do they seem to have (also probe on resources and who is providing them)? How are things coordinated?
3. How far along ('mature') do you think the work of the network is in your country? (Probe: is it still being developed? or would you say it is fully developed?)
4. What scale is the network operating at in your country? is it in a small set of focal areas? if so, how many hospitals and districts? or does it cover a larger scale? has it expanded or contracted since it started in your country? if so, how? and why?
5. [if at a network meeting:] Who is here at this meeting? Which stakeholders? Is there anyone important missing? why is each stakeholder you mention important for the network in your country?
6. Do you perceive the network activities in your country to be 'network' activities or part of broader quality improvement or maternal, newborn and child health efforts?
7. Does the work of the network in your country build on any previous efforts to improve the quality of care for mothers, newborns and children? if so, what were these previous initiatives? and what became of them?
8. Are there any other current initiatives in your country that may work well together with the work of this network? If so, what are they and what do they do? which stakeholders are involved in these initiatives and why are they important?
9. Are there any other initiatives in your country that may work against the work of this network? If so, what are they and what do they do? which stakeholders are involved in these initiatives and why might their agenda conflict with that of the network?

10. Does this network add value to efforts in your country to reduce maternal and newborn case fatality rates in health facilities? if so, how? if not, why not?
11. What does your country plan to do as a result of this network?
12. Has any of your recent work been influenced by the work of the network? if so, how?
13. Are there aspects of your country's health system that make it easier for the network to achieve its goals? if so, what are they? and why do they make it easier for the network to achieve its goals?
14. Are there aspects of your country's health system that make it more difficult for the network to achieve its goals? if so, what are they? and why do they make it more difficult for the network to achieve its goals?

##### The work of the network

15. What do you understand the goals of this network to be? what do you think of these goals?
16. What is your opinion of the work of this network?
17. Which aspects of the network do you like? and why do you like them?
18. Which aspects of the network could be improved? why do they need improving? and how might they be improved?
    a. Probe if not already mentioned: is there any conflict between 'central –global level- goals' and 'local –national and district level- goals?)
    b. Probe if not already mentioned: do you think this meeting is valuable? if so, why? if not why?

##### Your involvement in the network

19. How long have you been involved in the network and what is your role in the network? (Probe: what do you actually do as part of the network? what do you share with other countries? or the central coordinating team?)
20. Do you feel part of a network? and a country team?
21. Why are you here at this meeting?
22. What do you hope to achieve by participating in this meeting?
23. What do you plan to do as a result of this meeting?
24. Do you feel that your involvement in the network is shaping its agenda, direction or specific activities? if so how, and why? if not why?
25. Who do you think the most important partners are in the network at global level? and why?
26. Who do you think the most important partners are in the network at national level? and why?
27. Who have you shared the idea of the network with? (Probe: name 5 people who you have introduced to the network, their position and relation to you)

##### Appendix 2: First round local level interview topic guide

##### Interview topic guide for Quality of Care Network Hospital level stakeholders

*Information to read to interviewees first:* We are undertaking a research project on the quality of care network and how it operates. We would like to get the opinion of key stakeholders like yourself on how the network is doing. With your permission, we would like to ask you some questions on the work of the network at hospital level and in your country, and your personal role in the network and that of your team. This research project is funded by the UK Medical Research Council Health Systems Research Initiative and is being undertaken by Parent and Child Health Initiative in Malawi, Bangladesh Diabetic Association Perinatal Care Project in Bangladesh, Makerere University in Uganda, University College London, Oxford University and Johns Hopkins University, who are evaluating the work of the network as a whole. There are no right or wrong answers or any consequences (positive or negative) of taking part in this interview, and we would really like you to feel free to express your opinions without worrying about what they are. All information will be used to improve the work of the network and future networks. You are free to refuse to take part in this interview and to stop the interview at any time. With your permission we would like to record this interview so we can fully consider all of your responses. The recording and any notes I take will be kept secure and you will remain anonymous in any report stemming from this work.
Permission to be interviewed: signature: ______________________________________
Permission for interview to be recorded, signature:----____________________________________
Date: ___________________________

Which hospital are you representing? What is your position?

**The network and your hospital**

1. Tell me what you understand about how your hospital got involved in this network? Who seemed to really be leading efforts to join the network (probes: was it government, WHO, UNICEF, other organisations, or individuals?)
2. Who is part of the network now in your country? What roles do they seem to have (also probe on resources and who is providing them)? How are things coordinated?
3. How far along ('mature') do you think the work of the network is in your hospital (probe: is it still being developed? or would you say it is fully developed?)
4. What scale is the network operating at in your hospital? what activities happen at your hospital as a result of the network? how often do they happen? and when did these activities start? has the level of network activity increased or decreased since it started in your hospital? if so, how? and why?
5. How many network members do you have in your hospital? and how often do network members from your hospital meet? what do you discuss?
6. How often do you meet with network members from other hospitals? what do you discuss? and where do these meetings take place (probe: at your hospital, at their hospital, somewhere else?)
7. How often do you meet network members or leaders from national level? what do you discuss? and where do these meetings take place (probe: at your hospital, somewhere else?)
8. How often do you meet network members or leaders from other countries? (Probe: which countries and who?) what do you discuss? and where do these meetings take place (probe: at your hospital, somewhere else?)

9. Which stakeholders are involved in these network meetings involving your hospital? Are there important stakeholders missing? why is each stakeholder you mention important for the work of the network in your hospital?
10. Do you perceive the network activities in your hospital to be 'network' activities or part of broader quality improvement or maternal, newborn and child health efforts in your hospital?
11. Does the work of the network in your hospital build on any previous efforts to improve the quality of care for mothers, newborns and children? if so, what were these previous initiatives in your hospital? and what became of them?
12. Are there any other current initiatives in your hospital that may work well together with the work of this network? If so, what are they and what do they do? which stakeholders are involved in these initiatives and why are they important?
13. Are there any other initiatives in your hospital that may work against the work of this network? If so, what are they and what do they do? which stakeholders are involved in these initiatives and why might their agenda conflict with that of the network?
14. Does this network add value to efforts in your hospital to reduce maternal and newborn case fatality rates? if so, how? if not, why not?
15. What does your hospital plan to do as a result of this network?
16. Has any of your recent work been influenced by the work of the network? if so, how?
17. Are there aspects of your country's health system that make it easier for the network to achieve its goals? if so, what are they? and why do they make it easier for the network to achieve its goals?
18. Are there aspects of your country's health system that make it more difficult for the network to achieve its goals? if so, what are they? and why do they make it more difficult for the network to achieve its goals?

##### The work of the network

19. What do you understand the goals of this network to be? what do you think of these goals?
20. What is your opinion of the work of this network?
21. Which aspects of the network do you like? and why do you like them?
22. Which aspects of the network could be improved? why do they need improving? and how might they be improved?
    a. Probe if not already mentioned: is there any conflict between 'central –global level- goals' and 'local –national, district and hospital level- goals?)

##### Your involvement in the network

23. How long have you personally been involved in the network and what is your role in the network? (Probe: what do you actually do as part of the network? what do you share with other hospitals or the central coordinating team?)
24. Do you feel part of a network? and a hospital team?
25. Do you feel that your involvement in the network is shaping its agenda, direction or specific activities? if so how, and why? if not why?
26. Who do you think the most important partners organisations are in the network at national level? and why?
27. Who have you shared the idea of the network with? (Probe: name 5 people who you have introduced to the network, their position and relation to you)

#### Appendix 3: Second round global level interview topic guide

**Interview topic guide for Quality of Care Network Global level stakeholders**

*Information to read to interviewees first:* We are undertaking a research project on the quality of care network and how it operates. We would like to get the opinion of key stakeholders like yourself on how the network is doing. With your permission, we would like to ask you some questions on the work of the network at global level, and your personal role in the network and that of your team. This research project is funded by the UK Medical Research Council Health Systems Research Initiative and is being undertaken by Parent and Child Health Initiative in Malawi, Bangladesh Diabetic Association Perinatal Care Project in Bangladesh, Makerere University in Uganda, University College London, Oxford University and Johns Hopkins University, who are evaluating the work of the network as a whole. There are no right or wrong answers or any consequences (positive or negative) of taking part in this interview, and we would really like you to feel free to express your opinions without worrying about what they are. All information will be used to improve the work of the network and future networks. You are free to refuse to take part in this interview and to stop the interview at any time. With your permission we would like to record this interview so we can fully consider all of your responses. The recording and any notes I take will be kept secure and you will remain anonymous in any report stemming from this work.

Permission to be interviewed: signature: ______________________________
Permission for interview to be recorded, signature:----______________________________
Date: ____________________

Which organisation are you representing? What is your position?

**Your organisation and the network, and the future of the network**

1. What do you understand about the funding of the network? who is funding it? and for how long? what do you think the next few years might look like in terms of funding? (Probe: is BMGF still providing funding? If yes, until when? If no, any other funding alternatives? how long do you think the network will be externally funded?)
2. How sustainable do you think the network is? (Probe: domestic country funding of coordination efforts? and implementation efforts?)
    a. What do you think will happen to the network after 2022?
3. [for new interviewees only] What does your organisation contribute to the network? what roles does it play? what resources does it provide?
4. Since the global meeting in March 2019 has there been any changes in which organisations are part of the network at the global level? If so, what roles do these new organisations seem to have? and what resources are they providing? How are things coordinated?
    a. What is the process for adding new partners at the global level? Have any new partners joined since March 2019? (Probe: Has GFF gotten involved? If so, what is their role? What resources are they providing)?
5. How far along ('mature') do you think the work of the network is? (Probe: is it still being developed? or would you say it is fully developed? are there elements that have facilitated or hindered maturity of the network?)
    a. What QCN activities have taken place at the global level since the last meeting in Addis Ababa in March 2019? how have these activities helped the network? and what still needs to be done?
    b. How has Covid-19 impacted QCN implementation? How has it impacted ongoing QCN activities? How has it impacted quality of care overall? (If Covid-

19 has already been mentioned: are there any other ways the pandemic impacted the work of the QCN we haven't discussed?)

6. [to save time could only ask to WHO respondents]: What scale is the network operating at globally? how many countries are involved? how do you envisage this number to change going forward? are more countries due to join? or might some leave? why?
   a. What is the process for adding new partner countries? (Probe: How/when did Kenya join the network?)
   b. What is the role of observer countries? How are they distinguished from full partners in terms of involvement, inclusion criteria, etc.? Do you know if any of the observer countries (or any other countries) intend to join soon? might new countries join after 2022? if so, how?
7. At what scale does the network operate within each country? has this changed? if so, how? or might it change? why do you think these national-level changes in scale of the network have occurred /will occur?
8. How does the LALA framework function in terms of implementation and M&E? (Probe: do the indicators correspond to LALA?)
9. What are your thoughts on the M&E framework? (Probe: do you agree with the indicators? How are they weighted? Is there anything in there that shouldn't be? Anything missing?)
10. How would you evaluate the success of the QCN at the global level? is your organization involved in evaluation of the QCN? If so, how?
11. Do you perceive the network activities to be 'network' activities or part of broader quality improvement or maternal, newborn and child health efforts?
12. Are there any other initiatives that your organisation is involved in that may work well together with the work of this network? If so, what are they and what do they do? which stakeholders are involved in these initiatives and why are they important? (Probe: Does this network add value to efforts by your organisation to reduce maternal and newborn case fatality rates in health facilities? if so, how? if not, why not?)
13. Are there any other initiatives that your organisation is involved in that may work <u>against</u> the work of this network? If so, what are they and what do they do? which stakeholders are involved in these initiatives and <u>why might their agenda conflict with</u> that of the network?

#### The work of the network

14. According to the WHO 2021 report on the network, the network aims to achieve a 50% reduction in maternal, newborn and stillbirth case fatality rates by the end of 2022. Do you think this is achievable? if so, why? if not, why not?
    a. Has the pandemic changed your views of the feasibility of achieving the QCN goals? if so, how?
15. What is your opinion of the work of this network?
16. Which aspects of the network do you like? and why do you like them?
17. Which aspects of the network could be improved? why do they need improving? and how might they be improved?

#### Your involvement in the network

18. How long have you personally been involved in the network and what is your role in the network?
19. Do you feel that your involvement in the network is shaping its agenda, direction or specific activities? if so how, and why? if not why?
20. Who do you think the most important partners are in the network at global level? and why?
21. Who do you think the most important partners are in the network at national level? and why?
22. Who do you think we should also interview at the global level for this research on the QCN network?

Now that I've finished with my questions, is there is anything you'd like to add? or is there is anything we didn't touch upon that you think is important?

### Appendix 4 – Example template for facility observations

**Part 1: QCN Evaluation: Hospital observations**

This observation checklist should not be shown to those you are observing so as to avoid behaviour change due to observation.

Observations should be undertaken in the labour, delivery, and post-natal wards and neonatal unit after gaining verbal consent from the hospital in-charge following reading the information sheet for hospital observations to them and giving them a copy of the information sheet.

**Health facility**: Anaka Hospital
**Type of Health Facility**: Public Hospital
**Number of staff present on observation day**: 9
**Date of observation:** 18/11/2020
**Observer:** Gloria Seruwagi
**Observer position:** Co-Investigator

For each woman tick ✓ or write Y if YES and cross ✗ or write N if NO next to each of the behaviours below

**1 Respectful maternity care practices**

| **Provider actions during initial assessment:** | Woman Observed | | | | | | | | |
|---|---|---|---|---|---|---|---|---|---|
| | 1 | 2 | 3 | 4 | 5 | 6 | 7 | 8 | 9 |
| Greets client in a respectful manner | Y | | Y | Y | | | | | |
| Encourages client to have support person | Y | | N | N | | | | | |
| Explains procedures before proceeding | Y | | Y | Y | | | | | |
| Informs client of findings | N | | Y | Y | | | | | |
| Asks client if she has any questions | N | | Y | Y | | | | | |
| **Provider actions during labour:** | | | | | | | | | |
| Provider explains what will happen during labor to client | | N | | | N | Y | | | |
| Provider encourages client to consume food and fluids during labor | | N | | | N | N | | | |
| Provider encourages or assists client to ambulate and assume different labor positions | | Y | | | Y | Y | | | |
| Provider supports client in friendly way during labor | | Y | | | Y | Y | | | |
| Provider drapes client before delivery | | Y | | | Y | Y | | | |
| **Provider actions after labour:** | | | | | | | | | |
| Provider keeps mother & newborn in the same room after delivery | | Y | | | Y | Y | | | |
| Provider explains procedures to the mother & ask for consent before handling newborn | | Y | | | Y | N | | | |

| | | | | | | |
|---|---|---|---|---|---|---|
| Provider handles newborn gently or safely | | Y | | | Y | Y |
| Provider gives breastfeeding support | | Y | | | Y | Y |
| Provider gives postpartum/postnatal check to the mother and newborn before discharge | | Y | | | Y | Y |
| **Provider actions for women who come to the facility after delivering at home: (**None at observation time) | | | | | | |
| Greets client in a respectful manner | | | | | | |
| Explains procedures before proceeding | | | | | | |
| Informs client of findings | | | | | | |
| Asks client if she has any questions | | | | | | |
| Provider gives postpartum/postnatal check to the mother and newborn | | | | | | |

These indicators are adapted from: Rosen HE, Lynam PF, Carr C, et al. Direct observation of respectful maternity care in five countries: a cross-sectional study of health facilities in East and Southern Africa. *BMC Pregnancy Childbirth* 2015; **15**: 306 & Sacks E. Defining disrespect and abuse of newborns: a review of the evidence and an expanded typology of respectful maternity care. *Reproductive Health* 2017; **14**: 66.

**Use a fresh table for each patient observed in labour (if more suitable, you can answer Y for Yes and N for No)**

**2 Cleanliness**

| | **Always** | **Mostly** | **Sometimes** | **Never** | **Not observed** | **Example / Comments** |
|---|---|---|---|---|---|---|
| The bed appears to be clean | | Y | | | | |
| The staff wash their hands before touching the woman | | Y | | | | |
| The staff wash their hands when they are soiled | | Y | | | | |
| The staff wear a fresh pair of gloves for every internal examination | | Y | | | | |
| The newborn bed/incubator appears to be clean | | Y | | | | |
| The staff wash their hands before touching the newborn | | Y | | | | |

| | | |
|---|---|---|
| The bed appears to be clean | | Y |
| The staff wash their hands before touching the woman | | Y |
| The staff wash their hands when they are soiled | | Y |
| The staff wear a fresh pair of gloves for every internal examination | | Y |

| | | |
|---|---|---|
| The newborn bed/incubator appears to be clean | | Y |
| The staff wash their hands before touching the newborn | | Y |

| |
|---|
| The bed appears to be clean |
| The staff wash their hands before touching the woman |
| The staff wash their hands when they are soiled |
| The staff wear a fresh pair of gloves for every internal examination |
| The newborn bed/incubator appears to be clean |
| The staff wash their hands before touching the newborn |

| |
|---|
| The bed appears to be clean |
| The staff wash their hands before touching the woman |
| The staff wash their hands when they are soiled |
| The staff wear a fresh pair of gloves for every internal examination |
| The newborn bed/incubator appears to be clean |
| The staff wash their hands before touching the newborn |

| |
|---|
| The bed appears to be clean |
| The staff wash their hands before touching the woman |
| The staff wash their hands when they are soiled |
| The staff wear a fresh pair of gloves for every internal examination |
| The newborn bed/incubator appears to be clean |
| The staff wash their hands before touching the newborn |

**Part 2: Quality of care: facility observation template**

***(To be filled by the facility key informers)***

1. **Maternal and newborn health service offered by the facility**

| Type of service | Where it is provided | Time |
|---|---|---|
| Delivery | Labour suite | 12 – 9pm |
| NICU (newborn care) | NICU | 12 – 9pm |

2. **Record number of clients per service provided over the past 6 months the ward register**

| Type of service | Oct | Nov | Dec | Jan | Feb | Mar | Apr |
|---|---|---|---|---|---|---|---|
| Deliveries (normal) | | | | | | | |
| Deliveries (Caesarean) | | | | | | | |

**3. What is the average provider to client ratio for the facility in the labor ward?**

| During the day | During the night |
|---|---|
| Maternity 3:34<br>Hospital 1:>50 | 3:34 (beds) |

**4. Information on medical staff available at the facility**

| | Number of Staff in each cadre |
|---|---|
| Medical Officer | 6 |
| Assistant Medical Officer | 0 |
| Clinical officer | 4 |
| Medical Assistant | 0 |
| Nursing Officer | 10 |
| Assistant Nursing Officer | 0 |
| Public Health Nurse | 0 |
| State Registered Nurse | 30 |
| State Registered Nurse Midwife | 4 |
| Enrolled Nurse Midwife | 15 |
| Nurse Midwife Technician | 0 |
| Nurse Assistant | 3 |
| Community Midwives | 0 |
| Pharmacist | 1 |
| Pharmacy Assistant | 1 |
| Hospital Ombudsman | 2 |
| Other (Specify) | 30 (support staff) |
| Radiologist | 1 |
| Inventory manager | 1 |
| Orthopaedic officers | 2 |
| Accountant SAA | 1 |

**5. Information on medical staff available for maternal and newborn care**

| | Number of Staff in each cadre |
|---|---|
| Medical Officer | 6 |
| Assistant Medical Officer | 0 |
| Clinical officer | 4 |
| Medical Assistant | 0 |
| Nursing Officer | 10 |
| Assistant Nursing Officer | 0 |

| | |
|---|---|
| Public Health Nurse | 0 |
| State Registered Nurse | 30 |
| State Registered Nurse Midwife | 4 |
| Enrolled Nurse Midwife | 15 |
| Nurse Midwife Technician | 0 |
| Nurse Assistant | 3 |
| Community Midwives | 0 |
| Social Welfare Officer | 0 |
| Clinical Dentist | 1 |
| Dental Assistant | 1 |
| Pharmacist | 1 |
| Pharmacy Assistant (Dispenser) | 1 |
| Other (Specify) | |

6. **Record number of staff over the past 6 months allocated to maternal and newborn care**

| | 2019 | | | 2020 | | | |
|---|---|---|---|---|---|---|---|
| Cadre | May | June | July | Aug | Sept | Oct | Nov |
| Nursing Assistant | 1 | 1 | 1 | 1 | 1 | 1 | 1 |
| Registered Nurse Midwife | 1 | 1 | 1 | 1 | 1 | 1 | 1 |
| enrolled midwives | 12 | 12 | 12 | 12 | 12 | 12 | 12 |
| medical officers | 6 | 6 | 6 | 6 | 6 | 6 | 6 |
| Theatre team (Nurse, theatre assistant, anaesthetician) | | | | | | | |

7. What is the highest and minimum qualification of the staff located to maternal and newborn health?

| Highest | Minimum |
|---|---|
| Degree holder - Nurse/Midwife<br>Doctors - Master's degree (Ob/Gyn) | Certificate |

**SECTION B**

1. How are resources allocated?
   a. Duty Rota for maternity, pre-natal and post-natal
      - **Minimum of 3 per unit (maternity, labour and NICU)**

   b. Medicines and supplies (**Severe bleeding**, is enough oxytocic available? Are they able to detect **Pre-eclampsia?** Does the facility have enough magnesium sulfate?
      - **Yes, to All**
2. Quality improvements activities taking place at the facility. (Who, When where and how? – Obtain minutes if available)
   - **NICU ward; waste segregation, community engagement, WIT teams**
3. Quality of care network activities taking place at the facility. (Who, when where and how? – Obtain minutes if available)
   - **Journaling, partograph use, QI team meetings**
4. Hospital level meetings observations
   - **In charges of all units come together on a weekly, sometimes more frequent, basis to discuss key emerging issues.**
5. Ward level meetings observations

- **Observed mostly the interactions in the duty room, on ward and around the facility. Also, an informal meeting with the PNO and ADHO; cordial, supportive, improvement focused and supportive of facility priorities**.

6. How far is the nearest water supply from the beds?
   - **Approximately 20 metres (quite near)**

**Appendix 5 QCN Survey tool (adapted for use in each country)**

**Quality of Care Network Survey**

#### Preamble

You have been invited to participate in this study because you have been involved with The Network for Improving Quality of Care for Maternal, Newborn and Child Health.
This research project is funded by the UK Medical Research Council Health Systems Research Initiative. The research is being undertaken by Parent and Child Health Initiative in Malawi, Bangladesh Diabetic Association Perinatal Care Project in Bangladesh, Makerere University in Uganda, University College London, Oxford University and Johns Hopkins University, and aims to examine the determinants of successful clinical networks.
We are interested to find out what makes some networks more successful than others. The results from this study will inform the establishment and maintenance of clinical networks so they can effectively improve the quality of care.
Participation in this survey is entirely voluntary. Submitting a completed survey is an indication of your consent to participate in the study. You can withdraw from the study at any time. All aspects of the study, including the results, will be strictly confidential. Individuals will be de-identified in all reports relating to this study and will be labelled only with identification numbers. Network managers and chairs will not be identified by name in the publication of the results.
If you would like further information on the study and how your responses will be used, please read the participant information sheet.

#### Section 1 – Network membership

1. In what year did you join the network or start to be involved? ____________

1.2 What was/is your role in network?

- Chair/Executive Committee Member
- Executive & Steering Committee Member
- Expert Advisor
- Working group member
- Participant
-

- **Section 2 - Engagement** **The following questions are about the importance of the network to you:**

2.1 In the last 6 months how many hours have you devoted to network activities? E.g., attending network meetings, network correspondence, network quality improvement initiatives, network training activities

<1hr 1-5hrs 5-10hrs 10-20hr 20-30hrs
30-40hrs >40hrs

2.2 On the scale provided please rate the extent to which you agree or disagree with each statement:

| | strongly disagree | disagree | neither agree nor disagree | agree | strongly agree | Don't know |
|---|---|---|---|---|---|---|

| |
|---|
| I am committed to the network |
| I believe in the work that the network undertakes |
| I am not involved in the day-to-day work of the network |
| My input to the network is not highly visible but is more behind the scenes |
| My views and ideas have contributed to network activities |
| I have not been able to help drive the network agenda |

#### Section 3 - Clinical leadership

**The following questions relate to the leadership of the network**

3.1 Based on your personal experience, how much do you agree or disagree with each statement about the **Network Manager** **(name to be inserted)**

| | strongly disagree | disagree | neither agree nor disagree | agree | strongly agree | Don't know |
|---|---|---|---|---|---|---|
| The network manager had an evidence-based vision | | | | | | |
| The network manager was able to engage fellow professionals about service and quality improvement | | | | | | |
| The network manager brought others together to facilitate action and accomplish goals | | | | | | |
| The network manager built strong and positive relationships with clinicians | | | | | | |
| The network manager built strong and positive relationships with patients | | | | | | |
| The network manager built strong and positive relationships with hospital management | | | | | | |
| The network manager did not collaborate with external parties and administrators (e.g., WHO) to support network operations | | | | | | |

3.2 On the scale provided please rate the extent to which you agree or disagree with each statement about the collaborative work of the **Network Co-Chairs** **(names to be inserted)**:

| | strongly disagree | disagree | neither agree nor disagree | agree | strongly agree | Don't know |
|---|---|---|---|---|---|---|
| The network co-chairs did not make explicit the values and purpose of the network | | | | | | |

| |
|---|
| The network co-chairs were champions for change |
| The network co-chairs were not able to mobilize fellow professionals about service and quality improvement |
| The network co-chairs built strong and positive relationships with clinicians |
| The network co-chairs built strong and positive relationships with patients |
| The network co-chairs built strong and positive relationships hospital management |
| The network co-chairs did not collaborate with external parties and administrators (e.g., WHO) to support network operations |
| The network co-chairs worked cooperatively with senior leadership in WHO to make appropriate changes |

3.3 On the scale provided please rate the extent to which you agree or disagree with each statement about the role of the **Ministry of Health Quality Management Executive**

| | strongly disagree | disagree | neither agree nor disagree | agree | strongly agree | Don't know |
|---|---|---|---|---|---|---|
| The Ministry of Health Quality Management Executive provided strong leadership and clear strategic direction | | | | | | |
| The Ministry of Health Quality Management Executive worked cooperatively with the wider health system to make appropriate changes | | | | | | |

#### Section 4 - Internal management

**The following questions relate to how well you think the network was managed**

On the scale provided please rate the extent to which you agree or disagree with each statement about the management of the network:

| | strongly disagree | disagree | neither agree nor disagree | agree | strongly agree | Don't know |
|---|---|---|---|---|---|---|
| The network had multidisciplinary representation e.g., consumer, medical, nursing and allied health professionals | | | | | | |
| The network was dominated by a few individuals | | | | | | |
| The network provided a supportive environment allowing all voices to be heard | | | | | | |
| The network was effective in improving information sharing across the network | | | | | | |
| The network effectively coordinated communication with people and organizations outside the network | | | | | | |
| The network manager had good organisational abilities | | | | | | |

#### Section 5 - Perception of external support

**The following questions relate to the amount of support you believe the network received from external agencies or organisations**

5.1 On the scale provided please rate the extent to which you agree or disagree with each statement about the network's relationship with **Hospital Management**:

| | strongly disagree | disagree | neither agree nor disagree | agree | strongly agree | Don't know |
|---|---|---|---|---|---|---|
| There was strong support from hospital management for the work of my network | | | | | | |
| Hospital management were not willing to implement changes based on the recommendations of my network | | | | | | |
| Clinicians working in hospitals were willing to implement changes based on the recommendations of my network | | | | | | |

5.2 On the scale provided please rate the extent to which you agree or disagree with each statement about the networks relationship with **District Health Services**:

| | strongly | disagree | neither agree nor | agree | strongly | Don't know |
|---|---|---|---|---|---|---|

| | disagree | | disagree | | agree |
|---|---|---|---|---|---|
| District health managers were aware of the ideas put forward by my network | | | | | |
| District health managers were not willing to implement changes based on the recommendations of my network | | | | | |

5.3 On the scale provided please rate the extent to which you agree or disagree with each statement about the networks relationship with **national government**:

| | strongly disagree | disagree | neither agree nor disagree | agree | strongly agree | Don't know |
|---|---|---|---|---|---|---|
| The network workplans and agendas were aligned with government strategic plans | | | | | | |
| Government decision makers were not aware of the recommendations made by my network | | | | | | |

#### Section 6 - Perceived value

**The following questions relate to how much you believe the network has made a difference:**

Based on your experience, how much do you agree or disagree with each statement:

| | strongly disagree | disagree | neither agree nor disagree | agree | strongly agree | Don't know |
|---|---|---|---|---|---|---|
| The network's efforts have improved quality of care | | | | | | |
| The network's efforts have improved patient outcomes | | | | | | |
| The work of the network has not led to health system improvements | | | | | | |
| I would recommend joining this network to a colleague | | | | | | |
| The network does not help me professionally | | | | | | |

#### Section 7

**We may be developing some qualitative work in the future, exploring the features of successful networks... would you be interested in being asked to participate in this work that will involve an in-depth interview?**

YES NO

Do you have any additional comments you would like to make?

______________________________________________________________________________

#### Section 8 – About you

8.1 Gender:

- Male Female
-
- 8.2 What is/was your professional discipline (select ALL that apply):
- Medical Officer Nurse Patient Allied Health
- Executive manager - non-health professional Researcher/academic
- Other: ________________________________
-
- 8.3 Where was your primary place of work? (Select ONE):
- Hospital- principal referral
- Hospital – district
- Hospital – community
- Hospital – private
- Health Centre
- Other (specify) ______________________________________
-
-
-
-
-
- THANK YOU FOR YOUR TIME
